# Development and implementation of optimized endogenous contrast sequences for delineation in adaptive radiotherapy on a 1.5T MR-Linear-accelerator (MR-Linac): A prospective R-IDEAL Stage 0-2a quantitative/qualitative evaluation of *in vivo* site-specific quality-assurance using a 3D T2 fat-suppressed platform for head and neck cancer

**DOI:** 10.1101/2022.06.24.22276839

**Authors:** Travis C. Salzillo, M. Alex Dresner, Ashley Way, Kareem A. Wahid, Brigid A. McDonald, Sam Mulder, Mohamed A. Naser, Renjie He, Yao Ding, Alison Yoder, Sara Ahmed, Kelsey L. Corrigan, Gohar S. Manzar, Lauren Andring, Chelsea Pinnix, R. Jason Stafford, Abdallah S.R. Mohamed, John Christodouleas, Jihong Wang, Clifton David Fuller

## Abstract

**Purpose:** In order to improve segmentation accuracy in head and neck cancer (HNC) radiotherapy treatment planning for the 1.5T MR-Linac, 3D fat-suppressed T2-weighted MRI sequences were developed and optimized.

**Methods:** After initial testing of fat suppression techniques, SPectral Attenuated Inversion Recovery (SPAIR) was chosen as the fat suppression technique. Five candidate SPAIR sequences and a non-suppressed T2-weighted sequence were acquired on five HNC patients on the Unity MR-Linac. The primary tumor, metastatic lymph nodes, parotid glands, and pterygoid muscles were delineated by five segmentors. A robust image quality analysis platform was developed to objectively score the SPAIR sequences based on a combination of qualitative and quantitative metrics.

**Results:** Sequences were analyzed for signal-to-noise (SNR), contrast-to-noise (CNR) compared to fat and muscle, conspicuity, pairwise distance metrics, segmentor assessment, and MR physicist assessment. From this analysis, the non-suppressed sequence was inferior to each of the SPAIR sequences for the primary tumor, lymph nodes, and parotid glands, but was superior for the pterygoid muscles. Two SPAIR sequences consistently received the highest scores among the analysis categories and are recommended for use to Unity MR-Linac users for HNC radiotherapy treatment planning.

**Conclusions:** Two deliverables resulted from this study. First, an optimized 3D fat-suppressed T2-weighted sequence was developed that can be disseminated to Unity MR-Linac users. Second, a robust image quality analysis process pathway, used to objectively score the various SPAIR sequences, was developed and can be customized and generalized to any image quality optimization. Improved segmentation accuracy with the proposed SPAIR sequence can potentially lead to improved treatment outcomes and reduced toxicity by maximizing target coverage and minimizing organ-at-risk exposure.

## INTRODUCTION

Radiotherapy treatment planning using magnetic resonance imaging (MRI) exclusively, or at least in combination with computed tomography (CT), has become increasingly common over the past couple of decades [1]–[4]. The superior soft tissue contrast of MRI compared to CT makes it an attractive imaging modality for target structure and organ-at-risk (OAR) segmentation [5]–[7]. Furthermore, recent advances in deformable image registration and electron density assignment using synthetic CT generation or alternative atlas-based approaches, have helped address the primary pitfalls of combined MR/CT-based treatment planning—namely geometric distortion and direct dose estimation [8]–[16].

MR-Linac stakeholders are a major beneficiary of these advances in MR-based treatment planning [17], [18]. Hybrid MRI-linear accelerator devices can acquire a variety of imaging data during each fraction of radiotherapy and incorporate them into MR-compatible treatment planning systems [19], [20]. Moreover, these daily images can be used in on-line or off-line adaptive planning workflows when major changes in anatomy or tumor function are detected [21], [22]. Thus, a major area of research is in sequence development for these devices to better visualize relevant structures and discover and acquire useful imaging biomarkers for treatment response and resistance [23], [24].

Among the variety of tumor sites treated on MR-Linac devices, head and neck cancer (HNC), especially HPV-associated HNC, has demonstrated significant success on this device [25]–[27]. These tumors are relatively radiosensitive, which warrants the adaptive re-planning utility [28]–[30]. Furthermore, delineation of the complex anatomy in the head and neck region is difficult to visualize with CT, where most of the structures have uniform signal and little contrast; conversely, T2-weighted (T2w) MRI provides an ample amount of signal-to-noise as well as contrast with surrounding structures, which allows for clearer and more precise segmentation [31], [32].

However, when structures are adjacent to fat, which appears hyperintense on T2w MRI, the boundaries of various structures can become obfuscated [33], [34]. To attenuate the fat signal, while keeping the water signal within tissue intact, several fat-suppression methods have been established, which use pre-pulse inversion recovery and/or bandwidth strategies during image acquisition or post-processing techniques during image reconstruction. These fat suppression methods have been described at-length in the literature [35]–[40]. In the head and neck region, fat-adjacent structures are common, so a high-quality, fat-suppressed T2w MRI sequence is needed for the MR-Linac, which does not exist as of yet in published literature.

Thus, the purpose of this study was to develop and optimize a 3D fat-suppressed T2w sequence that could be used directly on a 1.5 T Unity MR-Linac for treatment planning purposes. Cast in the R-IDEAL (Radiotherapy-predicate studies, Idea, Development, Exploration, Assessment, Long-term study), as charged by the MR-Linac Consortium, this study was designed to represent Stage 0 (Radiotherapy Predicate Studies) to Stage 2a (Development) [41]. This provides a methodological and rigorous foundation for the implementation of this technical development for MR-Linac clinical workflows as well as the starting point for future studies along the R-IDEAL pipeline, which is an assessment methodology for evidence-based clinical evaluation of innovations in radiation oncology. The image quality analyses of the fat-suppressed sequence are presented here, along with the exam card of the optimized sequence itself, so that it may be disseminated to additional users of the Unity MR-Linac. As a secondary goal of this study, a comprehensive and robust image quality analysis platform was developed to objectively score and rank candidate fat-suppressed sequences. This analysis platform is easily customizable and can be generalized for the optimization of any anatomic-based imaging sequence.

## METHODS

### Data Availability

All patient images and segmentations were anonymized and uploaded to FigShare (DOI: 10.6084/m9.figshare.20140184).

### Sequence Development and Optimization

A standard 3D T2-weighted (T2w) turbo spin echo (TSE) sequence was utilized as an initial template for the fat-suppressed sequence. A Philips MR console emulation software (Philips Healthcare, Best, Netherlands) was used to modify sequence parameters and simulate relative image properties such as signal-to-noise (SNR). The first parameter that was iterated was the fat suppression method. Because there is no 3D mDixon sequence clinically available for the Unity device, only SPectral Attenuated Inversion Recovery (SPAIR) and Short Tau Inversion Recovery (STIR) techniques were investigated due to their relative resistance to B0 and B1 inhomogeneities that are known to occur in the neck region in MRI. Initial image acquisitions demonstrated that overall image quality was superior for the SPAIR fat suppression method compared to STIR, so subsequent sequence optimization was limited to SPAIR sequences. Several parameters were logically iterated (in contrast to spanning every possible combination in parameter space) to produce candidate SPAIR iterations which satisfied the following constraints: 5-6 minute acquisition time, ∼1 mm isotropic reconstructed resolution, TE and TR values for T2 weighting. A preliminary round of acquisition and qualitative analysis eliminated sequences that produced severe artifacts or insufficient image quality. Five SPAIR sequences were chosen as the final candidate sequences for further analysis. The parameters of these sequences can be found in Table 1.

**Table 1:** Relevant pulse sequence parameters for the non-suppressed T2-weighted sequence and candidate SPAIR sequences.

| Sequence Parameter | Non Fat Sat | SPAIR 1 | SPAIR 2 | SPAIR 3 | SPAIR 4 | SPAIR 5 |
| --- | --- | --- | --- | --- | --- | --- |
| Scan Mode | 3D | 3D |  |  |  |  |
| Technique | TSE | TSE |  |  |  |  |
| Fat Suppression | N/A | SPAIR |  |  |  |  |
| In-Plane FOV (mm) | 520 x 298 | 520 x 270 |  |  |  |  |
| In-Plane Acq. Resolution (mm) | 1.2 x 1.2 | 1.4 x 1.5 |  |  |  |  |
| Through-Plane FOV (mm) | 250 | 200 | 200 | 200 | 200 | 300 |
| Through-Plane Acq. Resolution (mm) | 2.2 | 2.0 | 2.0 | 2.0 | 2.0 | 2.4 |
| Reconstructed Voxel Size (mm) | 0.7 x 0.7 x 1.1 | 0.8 x 0.8 x 1.0 | 0.8 x 0.8 x 1.0 | 1.0 x 1.0 x 1.0 | 0.8 x 0.8 x 1.0 | 1.0 x 1.0 x 1.2 |
| Oversample Factor | 1.3 | 1.4 | 1.3 | 1.3 | 1.4 | 1.3 |
| TSE Factor | 150 | 72 | 76 | 66 | 72 | 76 |
| FID Reduction | Default | Strong | Through-Plane | Strong | Strong | Through-Plane |
| Flip Angle (°) | 90 | 90 |  |  |  |  |
| Refocusing Angle (°) | 30 | 40 | 40 | 40 | 55 | 40 |
| TE<sub>eff</sub> / TE<sub>equiv</sub> (ms) | 375 / 143 | 182 / 93 | 190 / 96 | 185 / 95 | 182 / 107 | 190 / 96 |
| TR (ms) | 2100 | 1600 | 1600 | 1400 | 1400 | 1400 |
| WFS (pix) / BW (Hz) | 0.473 / 459.3 | 0.407 / 533.4 | 0.456 / 476.6 | 0.498 / 436.4 | 0.407 / 533.4 | 0.459 / 473.3 |
| NSA | 2 | 2 |  |  |  |  |
| Scan Duration | 6:03 | 5:52 | 5:09 | 5:05 | 5:09 | 5:33 |

### Image Acquisition

Image data for the preliminary and main analysis were acquired on consenting head and neck cancer (HNC) patients who were enrolled in the MOMENTUM clinical trial at our institution (NCT04075305) and MD Anderson Institutional Review Board protocols PA15-0418 and PA18-0341. The images were acquired during both their MR Simulation and daily treatment fractions on the 1.5 T Unity MR-Linac device (Elekta AB, Stockholm, Sweden). The scanner is equipped with four-channel radiolucent RF coils positioned anteriorly and posteriorly to the patient, which is standard for Unity devices. A non-suppressed T2w and five SPAIR T2w sequences were each acquired on five HNC patients.

### Image Segmentation

Five post-graduate physicians in radiation oncology were asked to segment the following structures in each of the images using Raystation software (Raysearch Laboratories AB, Stockholm, Sweden). GTV (gross primary tumor volume), suspicious lymph nodes, left/right parotid glands, and left/right pterygoid muscles, which are relevant structures during radiotherapy treatment planning. The physicians (referred hereafter as segmentors) were restricted from looking at each other’s segmentations but were allowed to refer to a radiologist’s report for structure identification (which is a common clinical occurrence). Furthermore, the segmentors were asked to recontour each structure segmentation from scratch on each image, rather than propagating the segmentations onto each image and modifying them. A segmented structure on a particular sequence is referred hereafter as a structure-sequence pair. A non-resident researcher also segmented an air-filled cavity within the trachea in ten slices for each image. These were used for noise calculations since areas surrounding the patient were automatically masked in postprocessing before image export. Additionally, the non-resident researcher segmented three areas of cheek and neck fat on one slice for fat-CNR measurements.

### Quantitative Image Quality Analysis

#### Signal-to-noise and contrast-to-noise measurements

Signal-to-noise (SNR) of each structure was calculated as the mean signal of the sequence-structure pair divided by the standard deviation of the noise segmentation (ten slices of the air-filled cavity in the trachea). SNR was also calculated for the fat segmentations for each sequence as a measure of fat suppression.

Contrast-to-noise measurements between each structure and both fat and muscle were also determined by calculating the SNR difference between the structure and fat or muscle. The stack of SNR and CNR values for each sequence-structure pair was then combined for each segmentor and patient and used in the statistical analysis.

#### Conspicuity measurements

Conspicuity is a measurement of the ratio between ROI contrast and surrounding signal complexity. It is thought to be a more robust descriptor of structure visibility than SNR and CNR. A script to calculate conspicuity was developed according to the equations first described in [42] and is available at https://github.com/tcsalzillo/ConspicuityAnalysis. The original structure segmentations were isotropically expanded and contracted by 1 mm and 2 mm using Velocity AI software (Varian Medical Systems, Inc., Palo Alto, CA, USA). Because conspicuity was first formulated for 2D images, the conspicuity for each slice occupied by the structure for a particular sequence (sequence-structure pair) was recorded. The stack of conspicuity values for each sequence-structure pair was then combined for each segmentor and patient and inputted into the statistical analysis.

#### Pairwise Distance Metrics

Dice similarity coefficient (DSC) and 95% Hausdorff distance (HD) metrics for each sequence-structure pair between each segmentor were calculated as previously described [43]. These are amongst the most ubiquitous volumetric and surface distance metrics reported in literature [44]. The stacks of DSC and HD metrics for each sequence-structure pair were then combined for each segmentor and patient and inputted into the statistical analysis.

### Qualitative Image Quality Analysis

#### Segmentor Grading and Comments

As each segmentor worked to delineate the structures, they were asked to complete a rubric (Appendix 1) which asked the segmentor to qualitatively rank each sequence-structure pair, according to his/her preference, within each patient. Additionally, the segmentor was asked to provide specific comments about the appearance or visibility of a structure. These comments were classified into positive (e.g. Structure X looked great), neutral (e.g. Structure X looked acceptable), or negative (e.g. Could not see Structure X) categories. A metric was created to compare the relative amount of positive and negative comments for a specific sequence-structure pair, which was calculated with the following equation.

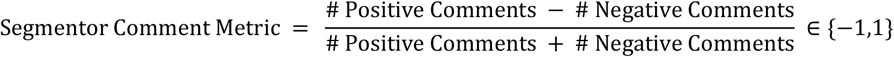

#### MR Physicist Assessment

Two MR physicists were asked to analyze the five SPAIR sequences according to a rubric (Appendix 2) which asked the physicist to qualitatively rank each sequence, according to his/her preference, within each patient. Additionally, the physicists were asked to identify any artifacts that were present in the image. One physicist quantified the number of slices that were affected by burnout (depletion of signal due to improper fat suppression) anteriorly and posterolaterally.

### Statistical Analysis

Each metric that was classified as quantitative (those in the SNR/CNR, Conspicuity, and Pairwise Distance categories) was subjected to further statistical analysis, which was performed using GraphPad Prism 8 (GraphPad Software, La Jolla, CA, USA). First, the distribution normality was assessed using the Kolmogorov-Smirnov test. If the distributions were found to be normal, the mean value and standard deviation of the metric were calculated. Statistical significance of the difference of means between each sequence-structure pair was then determined using the parametric one-way ANOVA test with follow-up Turkey multiple comparison corrections. If the distributions were found to be non-normal, the median value and interquartile range of the metric were calculated. Statistical significance of the difference of medians between each sequence-structure pair was then determined using the nonparametric Kruskal-Wallis test with follow-up Dunn’s multiple comparison corrections. In either case, statistical significance was attributed to comparisons that produced p<0.05.

### Rubric for Overall Sequence Scoring

For each metric that was analyzed, a score was determined for each sequence-structure pair (or just for each sequence if the metric was structure-agnostic such as in the MR Physicist Assessment). Each sequence-structure pair received a score between 1 and 6, where 6 corresponded to the pair with the best performance. For the metrics classified as quantitative, sequence-structure pairs could only receive a higher score than the other pairs if the difference was statistically significant (p<0.05) compared to all other sequence-structure pairs with a lower score. For example, if the SNR of Sequence A was statistically higher than B and C, then sequence A would be scored higher than B and C. However, if Sequence A was statistically higher than B but not C, and Sequence C was not statistically higher than B, then all 3 sequences would receive the same score. For metrics classified as qualitative, the statistical significance requirement was omitted, and sequence-structure pairs were scored purely according to their rank. Sequence-structure pairs that received the same score were rescaled to the average rank between them. For example, if a scoring distribution was scored as 6,5,5,5,5,1, it was rescaled to 6,3.5,3.5,3.5,3.5,1, where 3.5=(5+4+3+2)/4. For clarity, these will be regarded as Normalized Metric Scores

For each analysis category (SNR and CNR, Conspicuity, etc.), the Normalized Metric Scores within that category were summed for each sequence-structure pair. For metrics that are structure-agnostic, the score was added to each structure within the sequence. The summed score for a particular structure within a sequence was then renormalized between 1 and 6 (6 corresponding to highest summed score), according to its rank relative to the same structure among the other sequences. For clarity, these will be regarded as Normalized Category Scores. This normalization was performed so that a category with more metrics (such as SNR and CNR Measurements) would be weighed the same in the overall analysis as a category with fewer metrics (such as Conspicuity).

The Normalized Category Scores for each sequence-structure pair were then summed and normalized to determine the Total Score and Normalized Score. Initially, only the SNR and CNR, Conspicuity, Pairwise Distance, and Segmentor Analysis categories were summed since to compare the sequence-structure pairs between the non-suppressed sequence and SPAIR sequences. Then the MR Physicist Assessment Normalized Category Scores (only analyzed on the SPAIR sequences) were added to the Total Scores and renormalized to determine the Updated Total Scores and Updated Normalized Scores. These scores were used to compare the overall image quality for each structure among the SPAIR sequences. Additionally, the Updated Total Score for each structure within a sequence was summed and normalized to determine the Combined Total Score and Combined Normalized Score. These scores were used to compare the overall image quality across structures among the SPAIR sequences. Refer to Figure 1 for a graphical depiction of the scoring.

**Figure 1:**
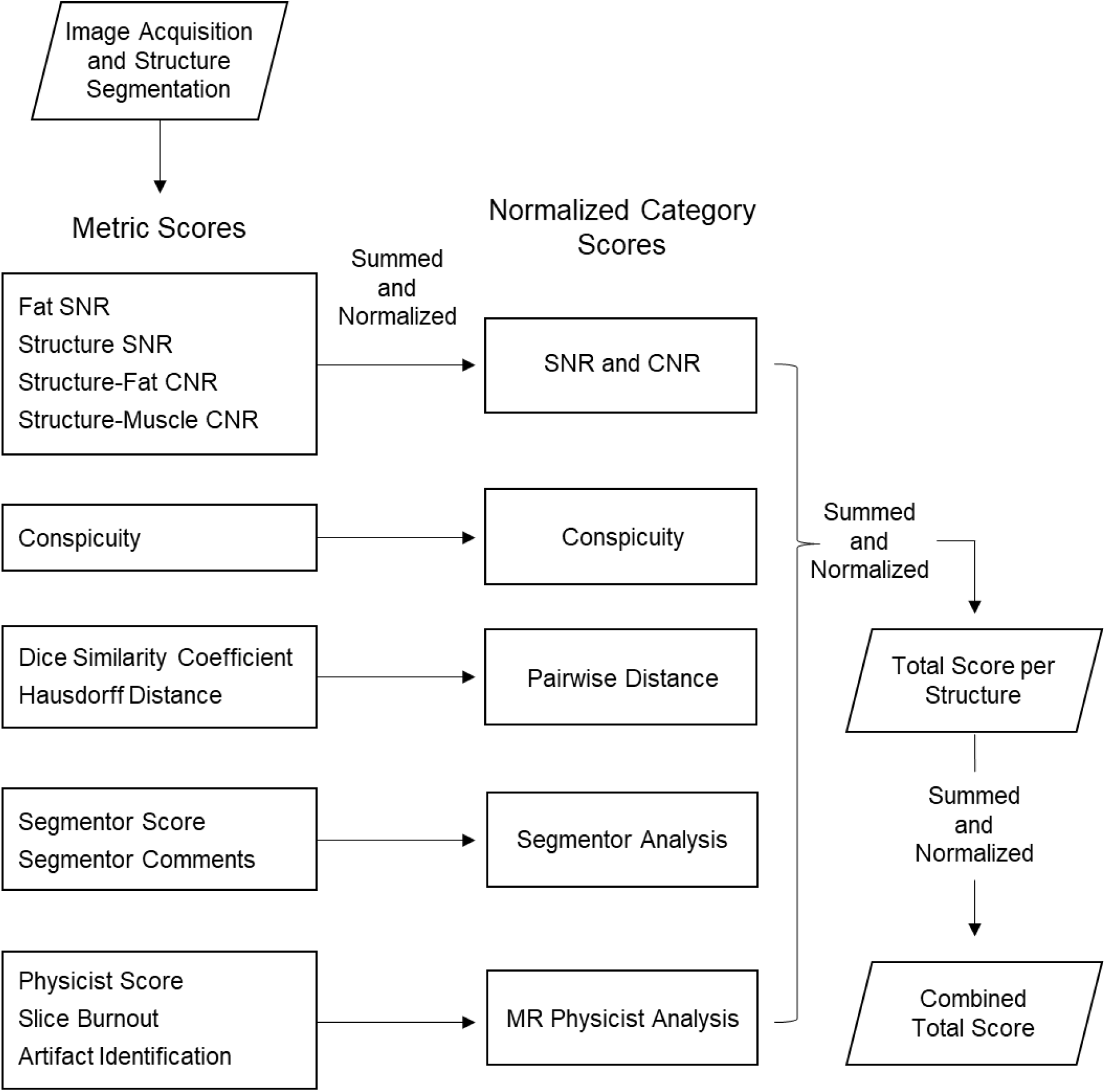
Flowchart of scoring rubric for the analysis platform. Metric Scores for each sequence-structure pair from each analysis are summed and normalized into their respective Normalized Category Scores. These Normalized Category Scores are further summed and normalized to calculate the Total Score for each sequence-structure pair. Lastly, the Total Score per structure within each sequence are summed and normalized to calculate the final Combined Total Score. During each “summed and normalized” step, weights can be applied if the user wishes to weigh an individual metric, individual analysis category, or individual structure higher for their specific application.

## RESULTS

### Image Acquisition and Segmentation

The non-suppressed T2w and each SPAIR sequence were successfully acquired on each of the five patients. Each of these images for a representative patient are shown in Figure S1. A representative pair of non-suppressed and SPAIR images (SPAIR 4) in two regions of the head and neck are illustrated in Figure 2 (with visible segmentations) and Figure S2 (without visible segmentations). The borders of the primary tumor and metastatic lymph nodes are clearer on the SPAIR image than the non-suppressed image. As a result, the segmentations, initially drawn on the non-suppressed image, clearly overestimate and underestimate the extent of the primary tumor and metastatic lymph nodes when viewed on the SPAIR image.

**Figure 2:**
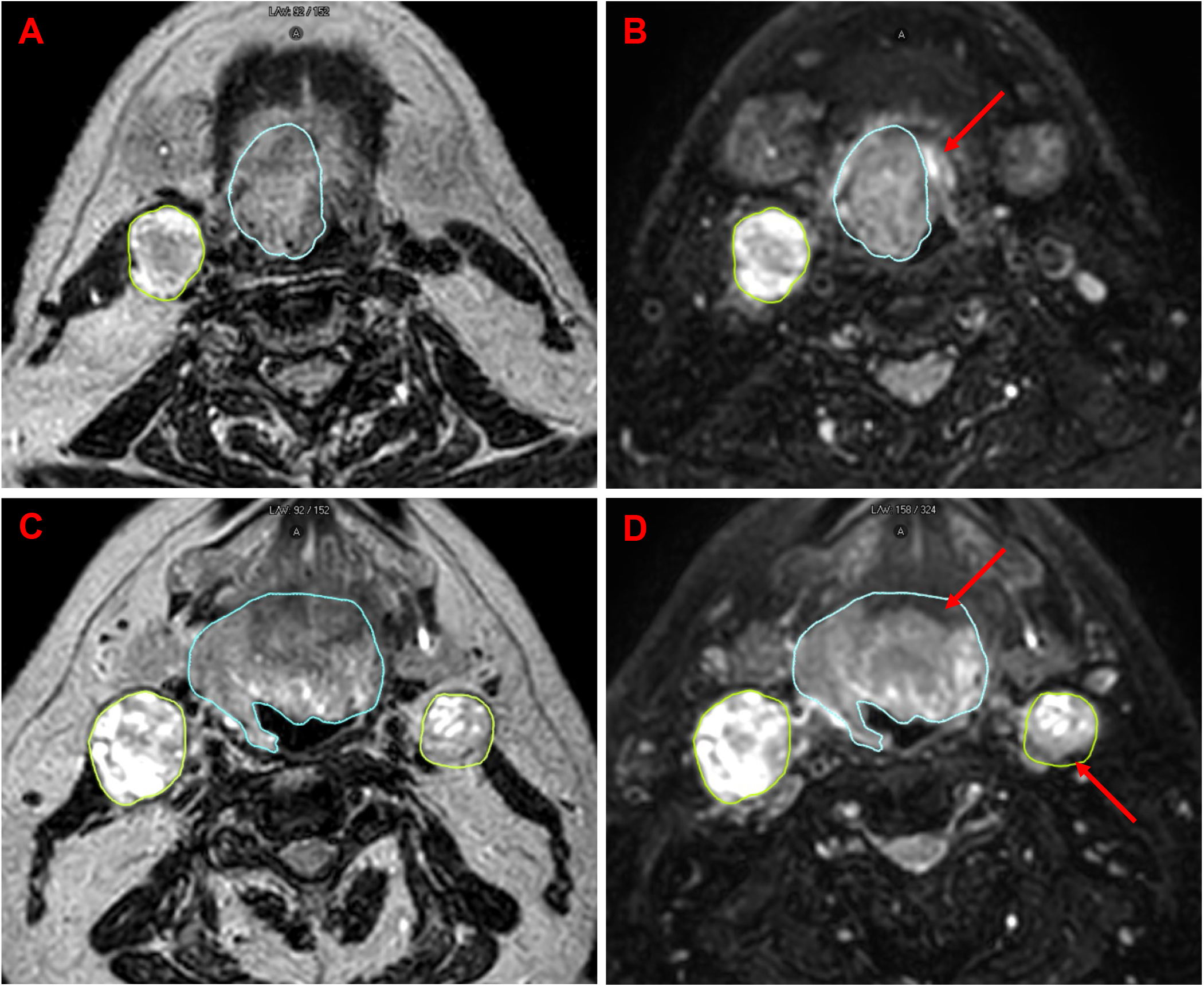
Representative cases of non-suppressed T2w (**A** and **C**) and SPAIR T2w (SPAIR4) (**B** and **D**) in a HNC patient. The top and bottom rows are two different slices in the image and illustrate the differences in primary and metastatic lymph node clarity. The visible segmentations were initially drawn on the non-suppressed T2w image. In the top row, the original segmentation underestimated the extent of the primary tumor, which is clearly visible on the SPAIR image (red arrow in **B**). The appearance of the inferior portion of the submandibular glands (bilateral structures anterior to the metastatic lymph node) can also be appreciated between the images; in the non-suppressed image, these glands are hypointense with little contrast to surrounding fat, but in the SPAIR image, these glands are hyperintense with exquisite contrast to the suppressed fat signal. In the bottom row, the original segmentations overestimated the extent of the primary tumor and lymph node, whose boundaries are better visualized on the SPAIR image (red arrows in **D**). Refer to Figure S2 to see the images without the segmentations visible.

### Quantitative Analyses

#### Signal-to-noise and contrast-to-noise measurements

Mean signal-to-noise (SNR) of segmented fat was significantly increased in the non-suppressed sequence compared to all other SPAIR sequences. Thus, each SPAIR sequence could effectively suppress fat signal. Among the SPAIR sequences, SPAIR 1 had the largest mean fat SNR of 3.1, which is still lower than the SNR of target and OAR structures. The SNR values for each structure were generally increased in the non-suppressed sequence, as expected, though it was only significantly increased the parotid glands compared to the SPAIR sequences.

Contrast-to-noise (CNR) between fat and the GTV/lymph nodes was significantly increased in the SPAIR sequences compared to the non-suppressed sequence. However, CNR between fat and the pterygoid muscles was significantly increased in the non-suppressed sequence compared to all SPAIR sequences. There were no significant differences in CNR between fat and the parotid glands among all sequences.

Furthermore, CNR between muscle and lymph nodes was significantly increased in the non-suppressed sequence and SPAIR 4 compared to all other SPAIR sequences, and CNR between muscle and parotid glands was significantly increased in the non-suppressed sequence compared to all SPAIR sequences. There were no significant differences in CNR between muscle and the GTV among all sequences. Refer to Table 2 for all SNR and CNR measurements.

**Table 2:** Signal-to-noise and contrast-to-noise measurements of the GTV, lymph nodes, parotid glands, and pterygoid muscles in the non-suppressed and SPAIR sequences. For each sequence, the signal-to-noise ratio (SNR) of fat was calculated, which quantifies the degree of fat suppression. For each structure within the sequences, the SNR was also calculated, along with the contrast-to-noise ratio (CNR) with relative to the fat and muscle signals. Values are presented as mean ± standard deviation. Values for a specific sequence-structure pair that are denoted with * are significantly greater (p<0.05) than all values for the structure in the column that are not annotated with *.

| Pulse Sequence | Fat SNR | Structure | Structure SNR | Structure-Fat CNR | Structure-Muscle CNR |
| --- | --- | --- | --- | --- | --- |
| Non Fat Sat | **20.6 ± 3.9\*** | GTV | 16.1 ± 4.9 | 4.5 ± 4.0 | 7.7 ± 4.4 |
|  |  | Lymph Nodes | 20.3 ± 5.8 | 3.2 ± 2.9 | **11.6 ± 5.3\*** |
|  |  | Parotid Glands | **15.9 ± 2.8\*** | 4.7 ± 2.1 | **7.6 ± 1.7\*** |
|  |  | Pterygoid Muscles | 8.5 ± 1.6 | **12.1 ± 2.2\*** |  |
| SPAIR 1 | 3.1 ± 1.1 | GTV | 16.5 ± 6.2 | **13.4 ± 5.4\*** | 7.7 ± 1.7 |
|  |  | Lymph Nodes | 12.5 ± 4.5 | **9.2 ± 4.2\*** | 5.9 ± 2.5 |
|  |  | Parotid Glands | 10.7 ± 4.6 | 7.5 ± 3.8 | 1.8 ± 1.1 |
|  |  | Pterygoid Muscles | 9.0 ± 4.4 | 5.9 ± 3.7 |  |
| SPAIR 2 | 2.4 ± 1.0 | GTV | 14.9 ± 5.3 | **12.5 ± 4.3\*** | 8.0 ± 2.3 |
|  |  | Lymph Nodes | 13.4 ± 6.7 | **10.5 ± 6.9\*** | 8.6 ± 3.3 |
|  |  | Parotid Glands | 9.3 ± 4.2 | 6.9 ± 3.3 | 2.4 ± 1.3 |
|  |  | Pterygoid Muscles | 7.1 ± 3.9 | 4.7 ± 3.1 |  |
| SPAIR 3 | 2.4 ± 0.8 | GTV | 13.7 ± 1.6 | **11.3 ± 1.0\*** | 7.3 ± 1.9 |
|  |  | Lymph Nodes | 10.1 ± 4.0 | **7.8 ± 4.4\*** | 6.2 ± 2.7 |
|  |  | Parotid Glands | 7.8 ± 1.6 | 5.5 ± 1.0 | 1.5 ± 0.8 |
|  |  | Pterygoid Muscles | 6.5 ± 2.2 | 4.1 ± 1.7 |  |
| SPAIR 4 | 2.8 ± 0.8 | GTV | 15.2 ± 4.0 | **12.5 ± 3.5\*** | 8.4 ± 2.5 |
|  |  | Lymph Nodes | 16.2 ± 7.0 | **13.3 ± 6.7\*** | **10.3 ± 3.8\*** |
|  |  | Parotid Glands | 9.4 ± 3.0 | 6.6 ± 2.6 | 2.5 ± 1.4 |
|  |  | Pterygoid Muscles | 7.0 ± 2.2 | 4.3 ± 1.9 |  |
| SPAIR 5 | 2.8 ± 0.6 | GTV | 14.0 ± 3.3 | **11.2 ± 2.8\*** | 7.1 ± 1.0 |
|  |  | Lymph Nodes | 11.7 ± 5.4 | **8.8 ± 5.1\*** | 6.1 ± 2.9 |
|  |  | Parotid Glands | 9.0 ± 3.1 | 6.2 ± 2.6 | 2.0 ± 1.1 |
|  |  | Pterygoid Muscles | 7.2 ± 2.6 | 4.4 ± 2.2 |  |

#### Conspicuity measurements

Conspicuity measurements for the structures in each sequence are depicted in Figure 3. Median conspicuity was significantly reduced in the non-suppressed sequence compared to one or more of the SPAIR sequences for the GTV, lymph nodes, and parotid glands. Conversely, conspicuity of the pterygoid muscles was significantly increased in the non-suppressed sequence compared to all SPAIR sequences. Among the SPAIR sequences, conspicuity of the GTV, parotid glands, and pterygoid muscles was significantly increased in SPAIR 1, 2, and 4 compared to SPAIR 3 and 5. Conspicuity of the lymph nodes was significantly increased in SPAIR 4 compared to all other SPAIR sequences.

**Figure 3:**
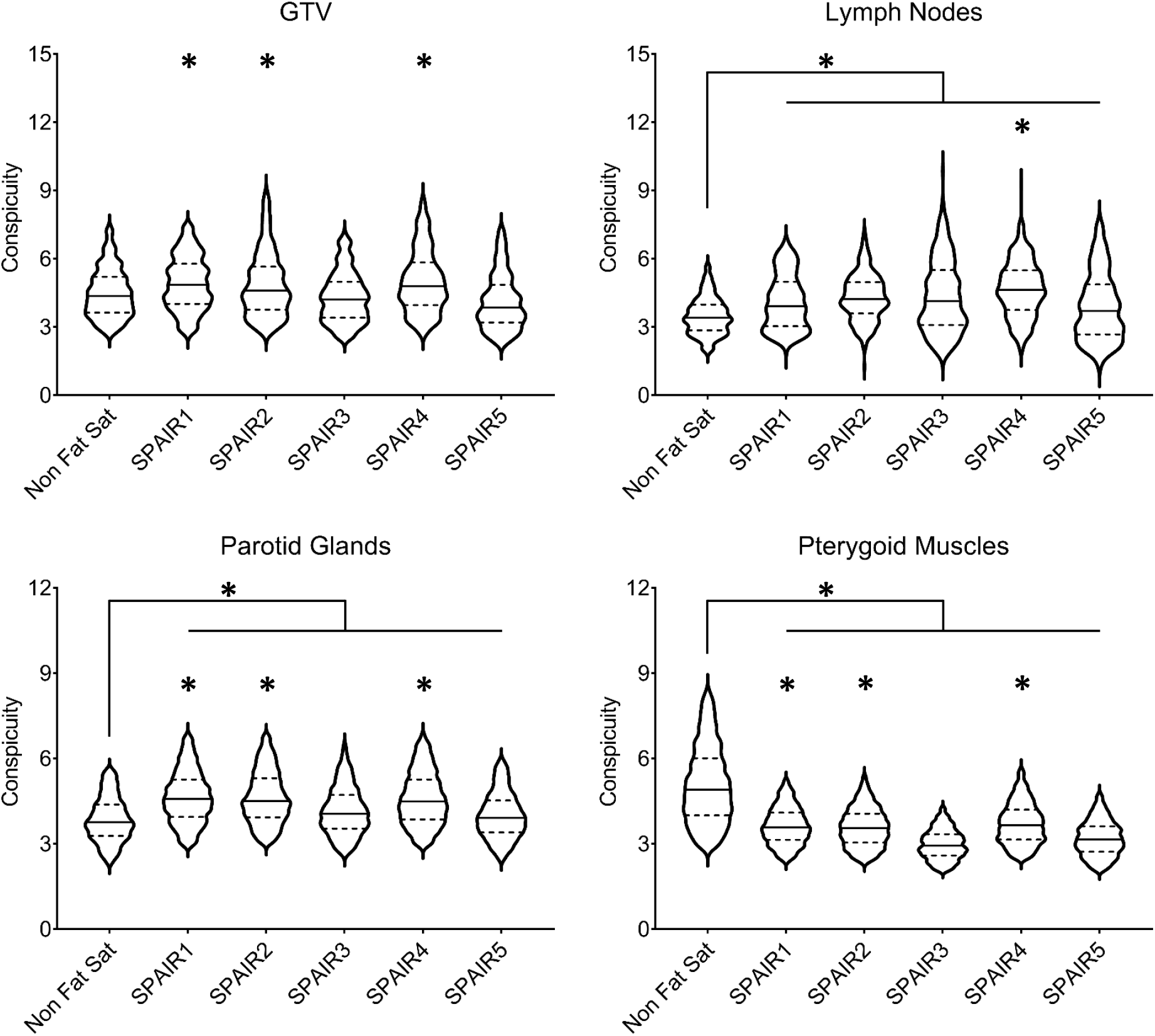
Conspicuity measurements of the GTV, lymph nodes, parotid glands, and pterygoid muscles in the non-suppressed and SPAIR sequences. Solid lines represent the median value of the distribution and dashed lines represent the limits of the interquartile range. Unless specifically annotated, all entries denoted with * are significantly greater (p<0.05) than all entries that are not depicted with *.

#### Pairwise Distance Metrics

The median of all pairwise dice similarity coefficient (DSC) measurements between each segmentor for the structures in each sequence are depicted in Figure 4. There was a large range of DSC values for the GTV in each sequence, and no significant differences were observed. DSC of the lymph nodes was significantly increased in SPAIR 2 and 4 compared to the non-suppressed image. DSC was significantly increased for the parotid glands and significantly decreased for the pterygoid muscles in all SPAIR sequences compared to the non-suppressed sequence.

**Figure 4:**
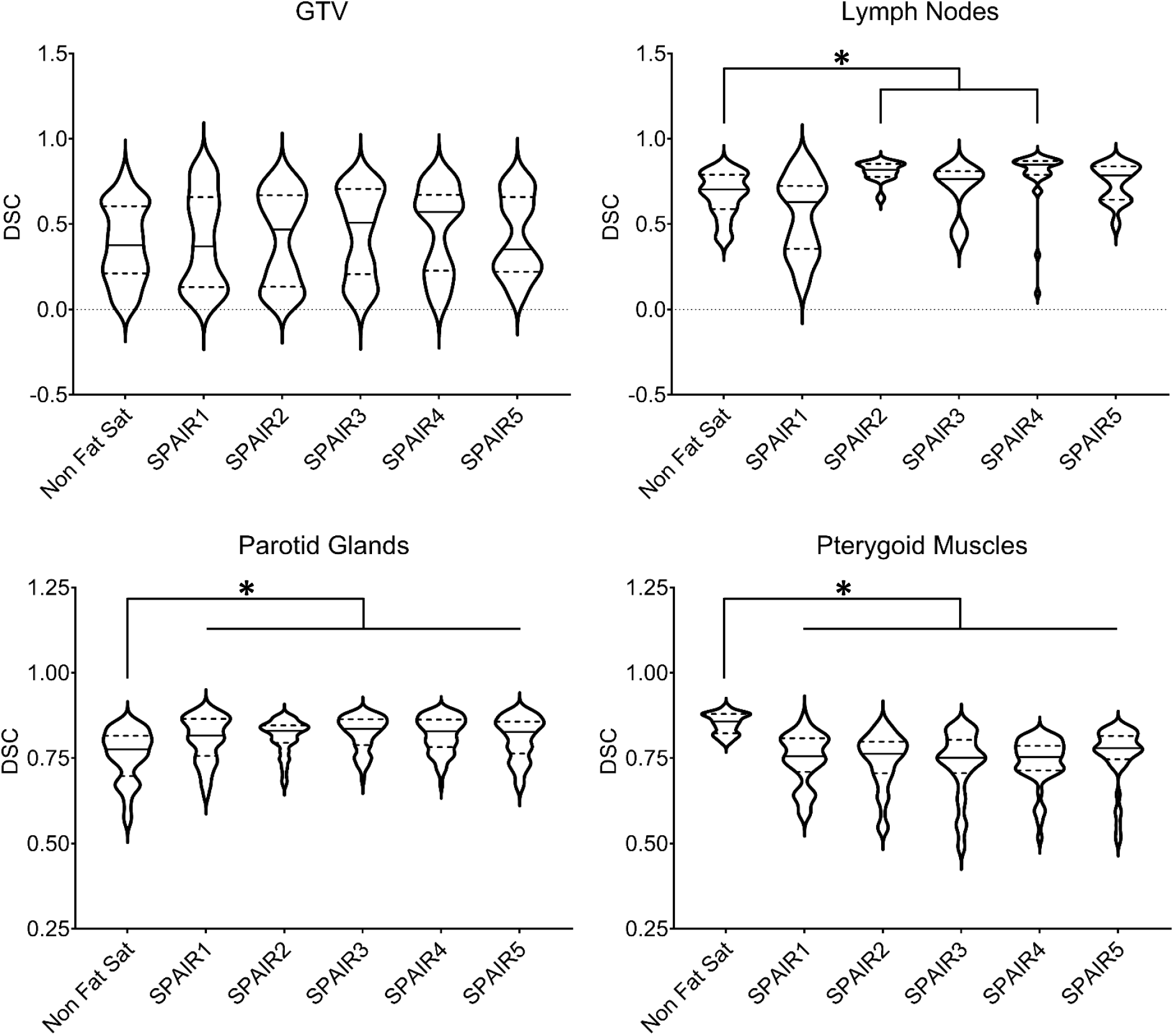
Dice similarity coefficient (DSC) measurements of the GTV, lymph nodes, parotid glands, and pterygoid muscles in the non-suppressed and SPAIR sequences. Solid lines represent the median value of the distribution and dashed lines represent the limits of the interquartile range. * indicates significant differences (p<0.05). Although some of the violin plot distributions extend below 0 and above 1, all DSC values were positive and between 0 and 1.

Hausdorff distance (HD) measurements followed a very similar trend as the DSC measurements and thus are illustrated in Figure S2. There was a large range of HD values for the GTV in each sequence, and no significant differences were observed. HD of the lymph nodes was significantly reduced in SPAIR 4 compared to the non-suppressed sequence. HD of the parotid glands was significantly reduced in SPAIR 2, 3, 4, and 5 compared to the non-suppressed sequence. Conversely, HD of the pterygoid muscles was significantly increased in all SPAIR sequences compared to the non-suppressed sequence.

### Qualitative Analyses

#### Segmentor Grading and Comments

Segmentor grades and number of positive and negative comments for each sequence are illustrated in Figure 5. SPAIR 1, 3, and 4 received the highest qualitative segmentor grades (higher grade is more preferred), though SPAIR 2 and 5 received only marginally worse grades. The non-suppressed sequence was consistently scored the lowest among the sequences. When looking at the relative number of positive and negative segmentor comments, only SPAIR 3 and 4 had net positive comments for all structures. The non-suppressed sequence had primarily negative comments for the GTV, lymph nodes, and parotid glands but primarily positive comments for the pterygoid muscles. SPAIR 5 had a relatively consistent number of positive and negative comments for each structure. Raw segmentor feedback is provided in Appendix 1.

**Figure 5:**
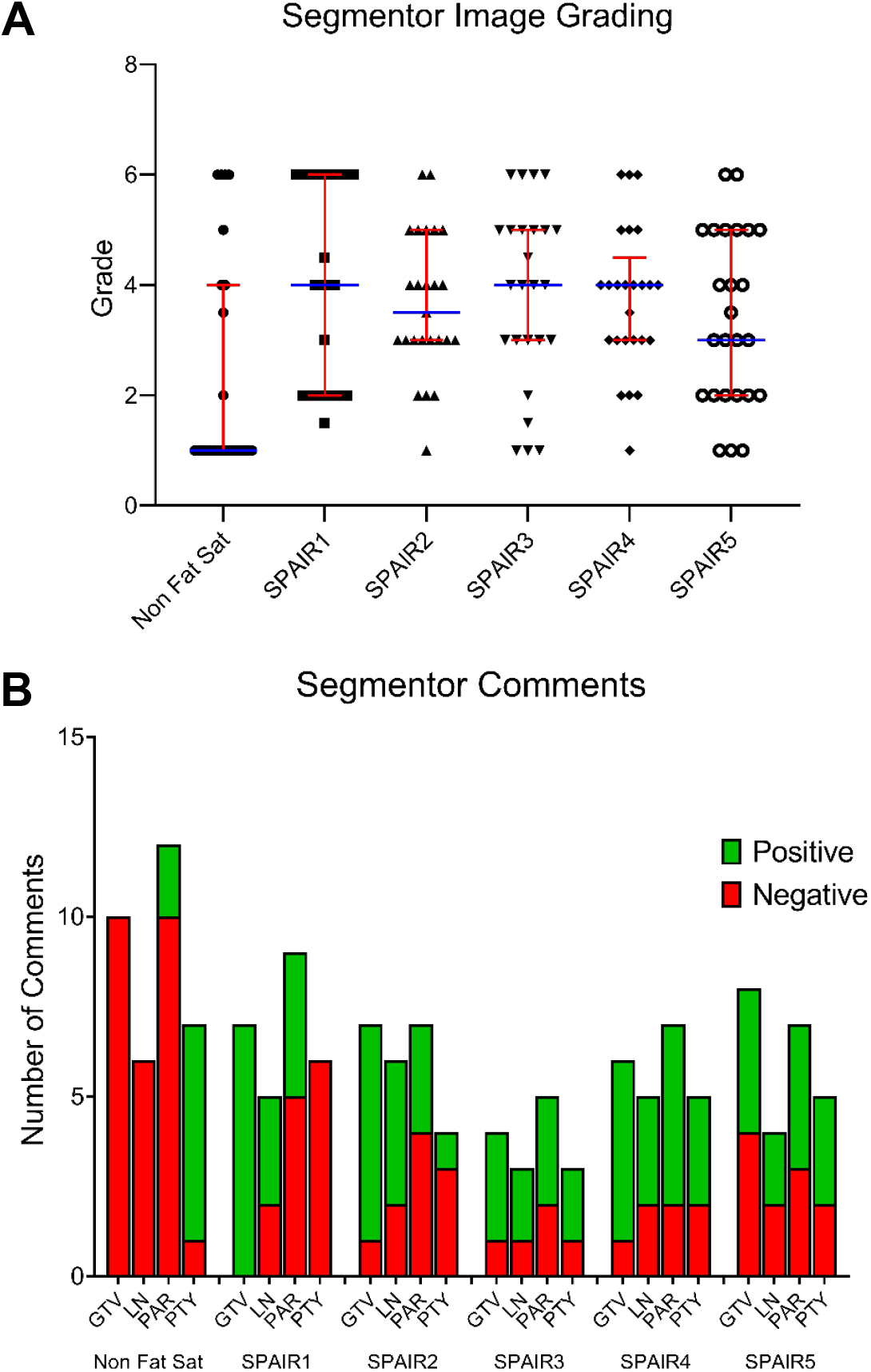
Segmentor image grading and comments of the GTV, lymph nodes, parotid glands, and pterygoid muscles in the non-suppressed and SPAIR sequences. In the plot of segmentor grades (**A**), median grade is depicted by the horizontal blue line, and interquartile range is depicted by the vertical red line. A higher grade corresponds to the more preferred sequence. The number of segmentor comments classified as positive and negative are illustrated in a stacked bar plot (**B**).

#### MR Physicist Assessment

MR physicists were asked to analyze only the SPAIR sequences, which is presented in Table 3. Among the SPAIR sequences, SPAIR 4 consistently received the highest qualitative physicist grades (most preferred) whereas SPAIR 2 and 5 consistently received the lowest grades. Moderate-to-severe artifacts were consistently observed in SPAIR 2 and 5, which included Herringbone (also called Filtering), Gibb’s Ringing, Zebra Artifact (also called 3D Phase Aliasing), and Partial Volume. One patient image acquired with SPAIR 4 did possess a Bright Blood Vessels artifact. On average, SPAIR 4 and 2 had the highest percentage of slices that displayed observable amounts of anterior and posterolateral burnout, respectively. Conversely, SPAIR 5 had the lowest percentage of slices that displayed observable amounts of anterior and posterolateral burnout. Raw MR physicist feedback is provided in Appendix 2. These artifacts are illustrated in Figures S4 and S5.

**Table 3:** MR physicists’ assessment of the non-suppressed and SPAIR sequences. A higher grade corresponds to the more preferred sequence. Numerical values are expressed as the median and interquartile range. Artifacts mentioned by either physicist on any patient image is listed.

| Pulse Sequence | Qualitative Score | Slices with Anterior Burnout | Slices with Posterolateral Burnout | Observed Artifacts |
| --- | --- | --- | --- | --- |
| SPAIR 1 | 4<br>(3.00 – 4.00) | 27.5%<br>(25.0 – 45.0%) | 17.5%<br>(15.0 – 25.0%) |  |
| SPAIR 2 | 2<br>(1.00 – 2.00) | 28.8%<br>(18.8 – 38.8%) | 25.0%<br>(22.5 – 30.0%) | Herringbone<br>Gibb's Ringing<br>Zebra Artifact |
| SPAIR 3 | 3.25<br>(3.00 – 4.25) | 27.5%<br>(21.3 – 37.5%) | 20.0%<br>(17.5 – 32.5%) |  |
| SPAIR 4 | 5<br>(4.75 – 5.00) | 32.5%<br>(20.6 – 35.0%) | 17.5%<br>(17.5 – 25.0%) | Bright Vessels |
| SPAIR 5 | 1.25<br>(1.00 – 2.00) | 24.0%<br>(15.0 – 34.0%) | 17.0%<br>(12.0 – 22.0%) | Herringbone<br>Gibb's Ringing<br>Partial Volume |

### Overall Sequence Scores

The results from the above analyses were analyzed according to the image analysis rubric by formulating them into Normalized Metric Scores for each structure-sequence pair and grouping into their respective categories (Tables S1-S5). Normalized Category Scores were then calculated. These Normalized Category Scores from the four categories that were analyzed for the non-suppressed and SPAIR sequences (SNR and CNR, Conspicuity, Pairwise Distance, and Segmentor Analysis), were summed and normalized (Table 4). The non-suppressed sequence accounted for the worst Total Score for the GTV, lymph nodes, and parotid glands, but the best Total Score for the pterygoid muscles. After incorporating the relative scores from the MR Physicist Assessment category for the SPAIR sequences, SPAIR 4 had the highest Updated Total Score for the lymph nodes, parotid glands, and pterygoid muscles, and the second highest for the GTV. SPAIR 1 had the highest Updated Total Score for the GTV and second highest for the lymph nodes, parotid glands, and pterygoid muscles. When considering all structures, SPAIR 4 had the highest Combined Total Score and SPAIR 1 had the second highest, though the scores for these sequences were very close.

**Table 4:** Overall scores for each sequence from each analysis category according to the image quality rubric. Normalized Category Scores for each structure from the SNR and CNR, Conspicuity, Pairwise Distance, and Segmentor categories were summed and normalized (6 = highest score) to calculate the Total and Normalized Score. Physicist scores, which were structure-agnostic and excluded the non-suppressed sequence, were then added to the Total Scores and renormalized to calculate the Updated Total and Normalized Scores. Lastly, the Updated Total Score for each structure within a sequence was summed and renormalized to generate the Combined Total and Normalized Scores for each sequence.

| Sequence | Non Fat Sat |  |  |  | SPAIR 1 |  |  |  | SPAIR 2 |  |  |  | SPAIR 3 |  |  |  | SPAIR 4 |  |  |  | SPAIR 5 |  |  |  |
| --- | --- | --- | --- | --- | --- | --- | --- | --- | --- | --- | --- | --- | --- | --- | --- | --- | --- | --- | --- | --- | --- | --- | --- | --- |
| Structure | GTV | LN | Par | Pty | GTV | LN | Par | Pty | GTV | LN | Par | Pty | GTV | LN | Par | Pty | GTV | LN | Par | Pty | GTV | LN | Par | Pty |
| SNR and CNR | 1 | 1 | 6 | 3.5 | 4 | 3.5 | 3 | 3.5 | 4 | 3.5 | 3 | 3.5 | 4 | 3.5 | 3 | 3.5 | 4 | 6 | 3 | 3.5 | 4 | 3.5 | 3 | 3.5 |
| Conspicuity | 2.5 | 1 | 1 | 6 | 5 | 3.5 | 5 | 4 | 5 | 3.5 | 5 | 4 | 2.5 | 3.5 | 3 | 1 | 5 | 6 | 5 | 4 | 1 | 3.5 | 2 | 2 |
| Pairwise Distance | 3.5 | 3.5 | 1 | 6 | 3.5 | 3.5 | 4 | 3 | 3.5 | 3.5 | 4 | 3 | 3.5 | 3.5 | 4 | 3 | 3.5 | 3.5 | 4 | 3 | 3.5 | 3.5 | 4 | 3 |
| Segmentor | 1 | 1 | 1 | 4 | 6 | 4 | 4 | 3 | 3.5 | 4 | 2 | 1 | 3.5 | 6 | 5 | 6 | 5 | 4 | 6 | 5 | 2 | 2 | 3 | 2 |
| Total Score | 8 | 6.5 | 9 | 19.5 | 18.5 | 15.5 | 16 | 13.5 | 16 | 15.5 | 14 | 11.5 | 13.5 | 16.5 | 15 | 13.5 | 17.5 | 19.5 | 18 | 15.5 | 10.5 | 12.5 | 12 | 10.5 |
| Normalized Score | 1 | 1 | 1 | 6 | 6 | 3.5 | 5 | 3.5 | 4 | 3.5 | 3 | 2 | 3 | 5 | 4 | 3.5 | 5 | 6 | 6 | 5 | 2 | 2 | 2 | 1 |
| MR Physicist |  |  |  |  | 6 |  |  |  | 2 |  |  |  | 4.5 |  |  |  | 4.5 |  |  |  | 3 |  |  |  |
| Updated Total Score |  |  |  |  | 24.5 | 21.5 | 22 | 19.5 | 18 | 17.5 | 16 | 13.5 | 18 | 21 | 19.5 | 18 | 22 | 24 | 22.5 | 20 | 13.5 | 15.5 | 15 | 13.5 |
| Updated Normalized Score |  |  |  |  | 6 | 5 | 5 | 5 | 3.5 | 3 | 3 | 2.5 | 3.5 | 4 | 4 | 4 | 5 | 6 | 6 | 6 | 2 | 2 | 2 | 2.5 |
| Combined Total Score |  |  |  |  | 87.5 |  |  |  | 65 |  |  |  | 76.5 |  |  |  | 88.5 |  |  |  | 57.5 |  |  |  |
| Combined Normalized Score |  |  |  |  | 5 |  |  |  | 3 |  |  |  | 4 |  |  |  | 6 |  |  |  | 2 |  |  |  |

## DISCUSSION

In this study, we developed several candidate SPAIR T2w sequences in order to incorporate fat suppressed images into the treatment planning pipeline for HNC patients treated on the Unity MR-Linac, which are reported here for the first time. We also developed a comprehensive image quality analysis platform to objectively score the sequences using a combination of quantitative and qualitative metrics. Using these metrics, the SPAIR sequence with the best combination of SNR, CNR, conspicuity, segmentation consistency, segmentor assessment, and MR physicist assessment was identified among four structures-primary tumor, metastatic lymph nodes, parotid glands, and pterygoid muscles. Both the optimized SPAIR sequence and the analysis platform can be utilized for clinical and research applications in radiation oncology.

The included metrics of the image analysis platform were carefully selected according to their applicability in HNC radiotherapy. For example, segmentation precision and qualitative comments were included in this analysis though they may not be as necessary for diagnostic imaging purposes. Furthermore, metrics for patient motion were not analyzed due to the use of immobilization devices in HNC radiotherapy. If optimizing sequences for thoracic or abdominal imaging, these metrics would need to be considered in optimal sequence selection. The major benefit of this image analysis platform is that it can be easily customized to add, remove, or weigh analysis metrics, depending on the application, and can be generalized to any sequence (MRI or other modality) for any number of structures. For example, if the user desired to weigh the results of the Segmentor and MR Physicist Assessments twice as much as the other analysis categories, they could simply double the Normalized Category Score of these analysis categories when calculating the Total Score or Updated Total Score. Alternatively, if they wanted the outcomes of the GTV and lymph node target structures to be considered twice as much as the parotid gland and pterygoid muscle OAR, they could simply double the scores of the former when calculating the Combined Total Score. We believe it is up to the individual user or institution to decide on how these weights should be implemented.

After combining the results from each individual analysis, excluding the MR Physicist Assessment (which was structure-agnostic and only performed on the SPAIR sequences), the non-suppressed sequence possessed lower scores than each SPAIR sequence for the GTV, lymph nodes, and parotid gland structures. Interestingly, the opposite was true for the pterygoid muscle structures, which received superior scores on the non-suppressed sequence compared to each SPAIR sequences. This was likely attributed to the fact that muscle appears hypointense in T2w sequences, so its contrast was further reduced after suppressing the signal from fat. Because the non-suppressed T2w sequences are characterized by improved contrast of these hypointense structures, generally higher SNR, and absence of fat-suppressed inhomogeneities and signal burnout relative to SPAIR sequences, we suggest that a non-suppressed T2w sequence should still be acquired for offline treatment planning purposes, rather than being completely replaced by a SPAIR T2w sequence. However, the superior performance of target structures in SPAIR sequences demonstrates their benefit in HNC radiotherapy treatment planning and are thus worth acquiring in addition to the non-suppressed T2w sequence.

After incorporating the MR Physicist Assessment for the SPAIR sequences, it was clear that SPAIR 1 and 4 emerged as the top performing sequences among the SPAIR iterations. While SPAIR 1 was slightly preferred for GTV segmentation, SPAIR 4 was slightly preferred for the remaining lymph node, parotid gland, and pterygoid muscle structures. When combining the scores for each structure, SPAIR 4 had a slightly better Combined Total Score than SPAIR 1. SPAIR 4 was also consistently scored as one of the top sequences for each analysis category among the SPAIR sequences. It should also be noted that the acquisition time for SPAIR 4 was nearly 1 minute less than SPAIR 1. The remaining parameters were very similar between the two sequences. The only differences were Refocusing Angle (40° vs. 55°), TE_equiv_ (93 vs. 107 ms), and TR (1600 vs. 1400 ms) for SPAIR 1 and 4, respectively. These sequences also had the largest Oversample Factor and smallest Water-Fat Shift compared to the remaining SPAIR sequences. Ultimately the user may want to test both sequences for their personal preference, but we officially recommend that SPAIR 4 be used for HNC radiotherapy treatment planning.

The two lowest scoring SPAIR sequences graded by the segmentors were also the two lowest graded by the MR physicists, which were the only sequences that produced numerous artifacts (SPAIR 2 and 5). Thus, these artifacts seem detrimental for radiation oncology applications, such as delineating target volumes and OAR. When looking at the parameters of these sequences, both used “Through-Plane” FID reduction instead of “Strong”. This parameter controls the crushing gradient that is responsible for attenuating residual magnetization during an echo train. Typically, this attenuation is achieved using the in-plane gradients (such as in the “Strong” option), but a research option for this parameter is “Through-Plane”, which uses the transverse gradient to attenuate the magnetization and reduces TE (also used in flow compensation). Insufficient magnetization attenuation can lead to stimulated echoes in an echo train and resultant “ringing-like” artifacts, which appear as alternating lines of hyperintense and hypointense signal in an image (example of this artifact and others shown in Figure S4, listed as Herringbone artifact). No artifacts were identified in any of the other sequences (except for “Bright Blood Vessels” in one of the SPAIR 4 patient images), which is encouraging for their use in radiation oncology applications. In each SPAIR sequence, there was some degree of burnout (loss of tissue signal) in areas near tissue-air or tissue-bone interfaces away from isocenter, but this was expected due to B0 inhomogeneity in these areas and did not impact segmentation.

Other than the absence of severe artifacts, additional criteria should be met to utilize MR images for radiation oncology applications. High geometric accuracy is arguably the most important to ensure accurate target coverage during treatment delivery. Geometric distortion on the Unity MR-Linac has been extensively investigated and reported to be approximately 1-2 mm for 350 mm diameter spherical volume (DSV), which still resulted in treatment plan accuracy within recommended tolerances in phantoms and patients [19], [45], [46]. We observed similar results in the non-suppressed and SPAIR 1 and 4 sequences using a geometric distortion phantom provided by Elekta (data provided in Table S6 and Figure S6). Furthermore, there were no significant differences in the distortion measurements between the non-suppressed sequence (currently used clinically) and either SPAIR sequence. Additionally, high resolution images are needed for accurate delineation of structures. Each of the SPAIR sequences had an isotropic reconstructed voxel size of 1 mm or less (except for SPAIR 5 which had a larger through-plane FOV and resultant voxel size). Lastly, the target structures and OAR need to be visible in the images for accurate and consistent segmentation. The SNR/CNR, Conspicuity, Pairwise Distance Metrics, and Segmentor analyses all demonstrate that the SPAIR sequences improve the detectability of representative structures compared to the non-suppressed T2w sequence.

There are a variety of fat suppression methods that have been established in MRI. These include Short Tau Inversion Recovery (STIR), Chemically Selective Saturation (CHESS), Spectral Presaturation with Inversion Recovery (SPIR), Spectral Attenuated Inversion Recovery (SPAIR), and Dixon techniques. The details of these techniques are well-described in the literature [35]–[40]. STIR suffers from lower SNR due to the attenuation of all tissue with the same T1 as fat but is less sensitive to B0 inhomogeneities. CHESS improves SNR by selectively attenuating fat signal but suffers significant effects of B0 and B1 inhomogeneities. SPIR is a hybrid of STIR and CHESS but suffers from the same B0 and B1 inhomogeneities as CHESS. SPAIR, similar to SPIR, selectively attenuates fat signal but does so using an adiabatic pulse, which helps offset the effects of B1 inhomogeneities at the expense of a higher specific absorption rate (SAR) in the patient. The Dixon technique results in moderate SNR, more robust fat suppression, and less sensitivity to both B0 *and* B1 inhomogeneities (with the help of postprocessing), though they also usually result in longer scan times due to acquisition of multiple images.

There have been several studies that evaluated one or more fat suppression methods for head and neck diagnostic imaging purposes [47]–[50]. A few studies further compared various fat-suppressed techniques in the head and neck region. Gaddikeri, et al. compared Dixon and STIR techniques in T2w images as well as Dixon and SPIR techniques in post-contrast T1w images [33]. The Dixon method outperformed both STIR and SPIR techniques when measuring signal intensity and reader-graded image quality. In a series of publications, Ma, et al. demonstrated that a triple echo Dixon technique was superior at suppressing fat signal to a higher degree and more uniformly in the head and neck region compared to CHESS and alternative Dixon techniques [34], [51]. These results were corroborated by Wendl, et al. [52]. Kawai, et al. compared coronal STIR and axial SPIR techniques for detecting metastatic lymph nodes in HNC [53]. The authors stated that both techniques performed comparably for detecting the metastases, though the STIR sequence was shorter and possessed fewer susceptibility artifacts compared to SPIR.

Our initial plan was to utilize Dixon fat suppression for further optimization. However, there are currently no mDixon sequences available to clinical users of the Unity MR-Linac, and only 2D mDixon sequences with T2-weighting are available to select research users. Two-dimensional sequences are generally unusable for treatment planning purposes for two reasons-the lack of 3D precision when delineating structures as well as the logistical inability to import 2D images into the treatment planning system. Because we wanted to develop and optimize a sequence that could be broadly disseminated to other Unity users without the need for a research patch, we opted not to pursue the development of a novel 3D T2w mDixon sequence. Thus, for the time-being, SPAIR is the optimal clinically available fat suppression technique for HNC treatment planning on the Unity MR-Linac. If 3D Dixon sequences are broadly enabled on the Unity, then future directions for this study are to compare an optimized 3D Dixon sequence with the optimized SPAIR sequence presented in this paper. It should be noted that in a study by Huijgen, et al., which compared Dixon and SPAIR sequences for musculoskeletal tumor imaging, there were minimal differences among several image quality metrics except for fat suppression homogeneity, which was superior in the Dixon sequences [54]. However, Dixon sequences did perform noticeably better in areas with large B0 inhomogeneities, which suggests that future investigation of Dixon sequences in HNC are warranted. Lastly, once compressed sensing is clinically available for the Unity MR-Linac, whose iterative reconstruction of undersampled data could accelerate sequences by up to 40%, it would be worthwhile to utilize the framework outlined in this paper to re-evaluate accelerated SPAIR sequences.

One limitation of this study was that only a few patients were included in the analysis, though for initial sequence development and optimization studies, this is not too unusual. The required time to manually segment multiple structures per image for multiple images per patient does limit the total number of patients that can realistically be included in one study. This limitation was mediated by including multiple independent segmentors. Further multi-institutional validation of these sequences within the MR-Linac Consortium would add power to these results. Furthermore, investigating the utility of these sequences for autosegmentation purposes would greatly increase the number of available segmentations. Another limitation to the study was that only front-end treatment planning effects (segmentation) were investigated, rather than downstream dosimetric implications. But because there was no ground-truth segmentation for these structures, only relative differences in downstream dose from differing segmentations would have been inferred. A study designed around the clinical implications of utilizing or excluding an optimized fat-suppressed sequence during the treatment planning process would be of great interest.

## CONCLUSIONS

A 3D SPAIR T2w sequence has been developed and optimized for HNC treatment planning on the Unity 1.5 T MR-Linac. This sequence, along with the remaining candidate sequences, are available for download for use on other Unity devices. An additional deliverable from this study is the robust image quality analysis platform that can be customized and generalized to any type of image optimization. This is the first study to report data on fat suppression methods in the head and neck using the Unity MR-Linac and provide a working sequence. Furthermore, this is the first study to report a SPAIR T2w image quality assessment for primary and metastatic structures in HNC. We believe that HNC treatment planning and subsequent treatment outcomes can be improved through the utilization of this sequence.

## Data Availability

All data produced are available online on FigShare (DOI: 10.6084/m9.figshare.20140184)

https://figshare.com/articles/dataset/Fat-suppressed_SPAIR_T2-weighted_MRI_sequences_for_head_and_neck_imaging_on_the_MR-Linac/20140184

