## Supplemental Data for "Development and implementation of optimized endogenous contrast sequences for delineation in adaptive radiotherapy on a 1.5T MR-Linear-accelerator (MR-Linac): A prospective R-IDEAL Stage 0-2a quantitative/qualitative evaluation of *in vivo* site-specific quality-assurance using a 3D T2 fat-suppresse"

| **Sequence** | **Non Fat Sat** | | | | **SPAIR 1** | | | | **SPAIR 2** | | | | **SPAIR 3** | | | | **SPAIR 4** | | | | **SPAIR 5** | | | |
|---|---|---|---|---|---|---|---|---|---|---|---|---|---|---|---|---|---|---|---|---|---|---|---|---|
| **Structure** | GTV | LN | Par | Pty | GTV | LN | Par | Pty | GTV | LN | Par | Pty | GTV | LN | Par | Pty | GTV | LN | Par | Pty | GTV | LN | Par | Pty |
| **Fat SNR** | 1 | | | | 4 | | | | 4 | | | | 4 | | | | 4 | | | | 4 | | | |
| **Structure SNR** | 3.5 | 3.5 | 6 | 3.5 | 3.5 | 3.5 | 3 | 3.5 | 3.5 | 3.5 | 3 | 3.5 | 3.5 | 3.5 | 3 | 3.5 | 3.5 | 3.5 | 3 | 3.5 | 3.5 | 3.5 | 3 | 3.5 |
| **CNR-Fat** | 1 | 1 | 3.5 | 6 | 4 | 4 | 3.5 | 3 | 4 | 4 | 3.5 | 3 | 4 | 4 | 3.5 | 3 | 4 | 4 | 3.5 | 3 | 4 | 4 | 3.5 | 3 |
| **CNR-Mus** | 3.5 | 5.5 | 6 | - | 3.5 | 2.5 | 3 | - | 3.5 | 2.5 | 3 | - | 3.5 | 2.5 | 3 | - | 3.5 | 5.5 | 3 | - | 3.5 | 2.5 | 3 | - |
| **Total Score** | 9 | 11 | 16.5 | 10.5 | 15 | 14 | 13.5 | 10.5 | 15 | 14 | 13.5 | 10.5 | 15 | 14 | 13.5 | 10.5 | 15 | 17 | 13.5 | 10.5 | 15 | 14 | 13.5 | 10.5 |
| **Normalized Score** | 1 | 1 | 6 | 3.5 | 4 | 3.5 | 3 | 3.5 | 4 | 3.5 | 3 | 3.5 | 4 | 3.5 | 3 | 3.5 | 4 | 6 | 3 | 3.5 | 4 | 3.5 | 3 | 3.5 |

Table S1: Signal-to-noise and contrast-to-noise Metric Scores

| Sequence | Non Fat Sat | | | | SPAIR 1 | | | | SPAIR 2 | | | | SPAIR 3 | | | | SPAIR 4 | | | | SPAIR 5 | | | |
|---|---|---|---|---|---|---|---|---|---|---|---|---|---|---|---|---|---|---|---|---|---|---|---|---|
| **Structure** | GTV | LN | Par | Pty | GTV | LN | Par | Pty | GTV | LN | Par | Pty | GTV | LN | Par | Pty | GTV | LN | Par | Pty | GTV | LN | Par | Pty |
| **Conspicuity** | 2.5 | 1 | 1 | 6 | 5 | 3.5 | 5 | 4 | 5 | 3.5 | 5 | 4 | 2.5 | 3.5 | 3 | 1 | 5 | 6 | 5 | 4 | 1 | 3.5 | 2 | 2 |
| **Total Score** | 2.5 | 1 | 1 | 6 | 5 | 3.5 | 5 | 4 | 5 | 3.5 | 5 | 4 | 2.5 | 3.5 | 3 | 1 | 5 | 6 | 5 | 4 | 1 | 3.5 | 2 | 2 |
| **Normalized Score** | 2.5 | 1 | 1 | 6 | 5 | 3.5 | 5 | 4 | 5 | 3.5 | 5 | 4 | 2.5 | 3.5 | 3 | 1 | 5 | 6 | 5 | 4 | 1 | 3.5 | 2 | 2 |

Table S2: Conspicuity Metric Scores

| Sequence | Non Fat Sat | | | | SPAIR 1 | | | | SPAIR 2 | | | | SPAIR 3 | | | | SPAIR 4 | | | | SPAIR 5 | | | |
|---|---|---|---|---|---|---|---|---|---|---|---|---|---|---|---|---|---|---|---|---|---|---|---|---|
| **Structure** | GTV | LN | Par | Pty | GTV | LN | Par | Pty | GTV | LN | Par | Pty | GTV | LN | Par | Pty | GTV | LN | Par | Pty | GTV | LN | Par | Pty |
| **DSC** | 3.5 | 3.5 | 1 | 6 | 3.5 | 3.5 | 4 | 3 | 3.5 | 3.5 | 4 | 3 | 3.5 | 3.5 | 4 | 3 | 3.5 | 3.5 | 4 | 3 | 3.5 | 3.5 | 4 | 3 |
| **HD** | 3.5 | 3.5 | 3.5 | 6 | 3.5 | 3.5 | 3.5 | 3 | 3.5 | 3.5 | 3.5 | 3 | 3.5 | 3.5 | 3.5 | 3 | 3.5 | 3.5 | 3.5 | 3 | 3.5 | 3.5 | 3.5 | 3 |
| **Total Score** | 7 | 7 | 4.5 | 12 | 7 | 7 | 7.5 | 6 | 7 | 7 | 7.5 | 6 | 7 | 7 | 7.5 | 6 | 7 | 7 | 7.5 | 6 | 7 | 7 | 7.5 | 6 |
| **Normalized Score** | 3.5 | 3.5 | 1 | 6 | 3.5 | 3.5 | 4 | 3 | 3.5 | 3.5 | 4 | 3 | 3.5 | 3.5 | 4 | 3 | 3.5 | 3.5 | 4 | 3 | 3.5 | 3.5 | 4 | 3 |

Table S3: Pairwise distance Metric Scores

| Sequence | Non Fat Sat | | | | SPAIR 1 | | | | SPAIR 2 | | | | SPAIR 3 | | | | SPAIR 4 | | | | SPAIR 5 | | | |
|---|---|---|---|---|---|---|---|---|---|---|---|---|---|---|---|---|---|---|---|---|---|---|---|---|
| **Structure** | GTV | LN | Par | Pty | GTV | LN | Par | Pty | GTV | LN | Par | Pty | GTV | LN | Par | Pty | GTV | LN | Par | Pty | GTV | LN | Par | Pty |
| **Segmentor Score** | 1 | | | | 5 | | | | 3 | | | | 5 | | | | 5 | | | | 2 | | | |
| **Segmentor Comment** | 1 | 1 | 1 | 6 | 6 | 3.5 | 3 | 1 | 5 | 5.5 | 2 | 2 | 3 | 5.5 | 5 | 5 | 4 | 3.5 | 6 | 3.5 | 2 | 2 | 4 | 3.5 |
| **Total Score** | 2 | 2 | 2 | 7 | 11 | 8.5 | 8 | 6 | 8 | 8.5 | 5 | 5 | 8 | 10.5 | 10 | 10 | 9 | 8.5 | 11 | 8.5 | 4 | 4 | 6 | 5.5 |
| **Normalized Score** | 1 | 1 | 1 | 4 | 6 | 4 | 4 | 3 | 3.5 | 4 | 2 | 1 | 3.5 | 6 | 5 | 6 | 5 | 4 | 6 | 5 | 2 | 2 | 3 | 2 |

Table S4: Segmentor grading and comments Metric Scores

| Sequence | SPAIR 1 | SPAIR 2 | SPAIR 3 | SPAIR 4 | SPAIR 5 |
|---|---|---|---|---|---|
| **Physicist Score** | 5 | 3 | 4 | 6 | 2 |
| **Slice Burnout** | 5 | 2 | 4 | 3 | 6 |
| **Artifacts** | 5.5 | 2.5 | 5.5 | 4 | 2.5 |
| **Total Score** | 15.5 | 7.5 | 13.5 | 13.5 | 10.5 |
| **Normalized Score** | 6 | 2 | 4.5 | 4.5 | 3 |

Table S5: MR physicist assessment Metric Scores

| Sequence | Non Fat Sat | SPAIR 1 | SPAIR 4 |
|---|---|---|---|
| **ML Distortion (mm)** | 1.03 ± 0.58 | 1.60 ± 0.56 | 1.65 ± 0.60 |
| **AP Distortion (mm)** | 0.90 ± 0.43 | 0.72 ± 0.31 | 0.70 ± 0.30 |
| **SI Distortion (mm)** | 1.11 ± 0.40 | 1.01 ± 0.37 | 0.97 ± 0.37 |

Table S6: Geometric distortion measurements along each anatomic plane measured at 300 mm diameter spherical volume (DSV). Measurements were made using the Philips geometric distortion phantom provided with the Unity MR-Linac. These values represent the $98^{th}$ percentile ± standard deviation and are less than 2 mm, which demonstrates no significant geometric distortion among the SPAIR sequences. When comparing the distributions using one-way ANOVA, there were no statistically significant differences between the SPAIR sequences, nor between either SPAIR sequence and the non-suppressed T2 sequence, which is conventionally used in the clinic. Refer to Figure S5 for an illustration of the distortion distributions along each anatomic plane.

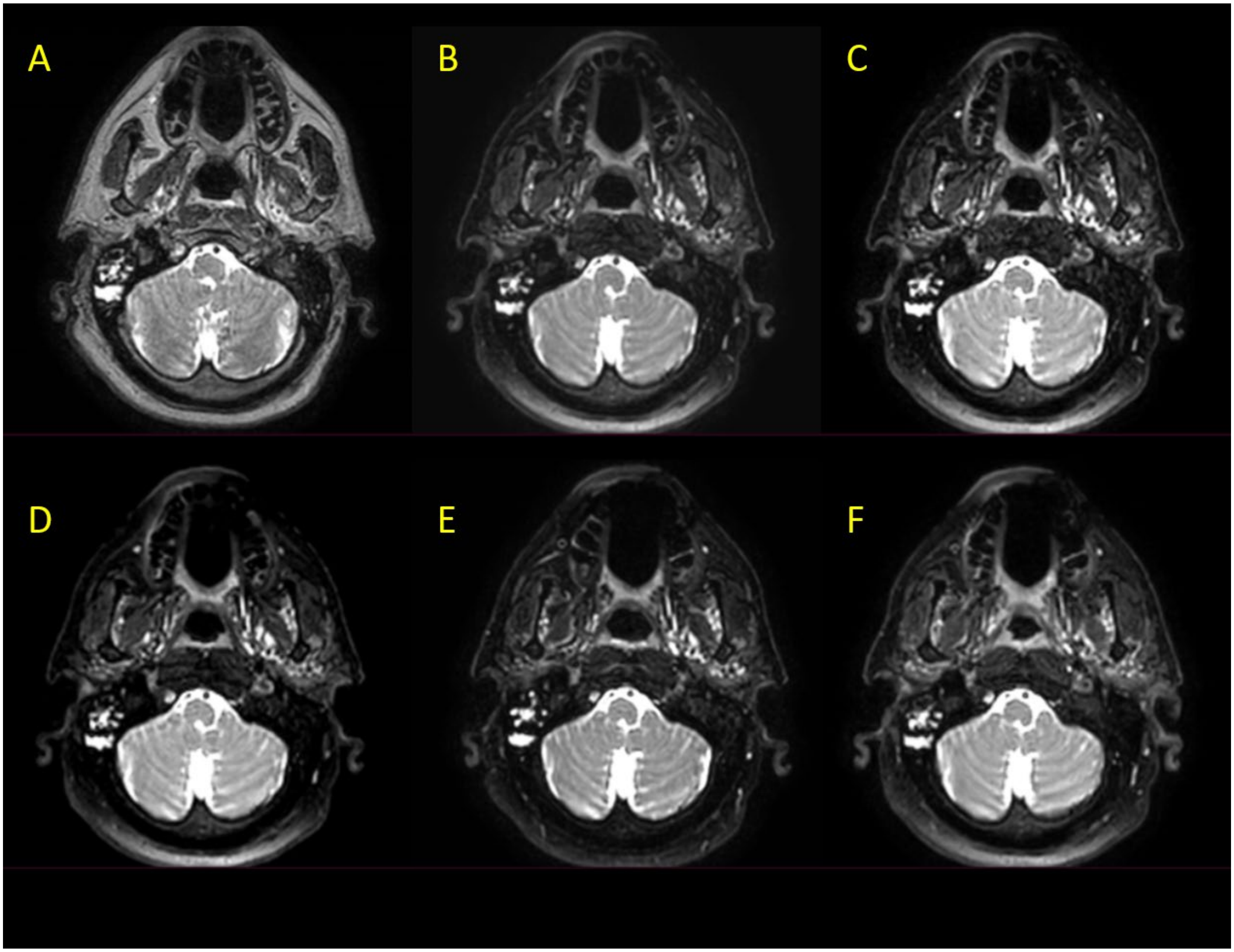


Figure S1: Non-suppressed T2w (**A**) and five SPAIR sequences (**B**-**F**) acquired on a representative HNC patient on the Unity MR-Linac.

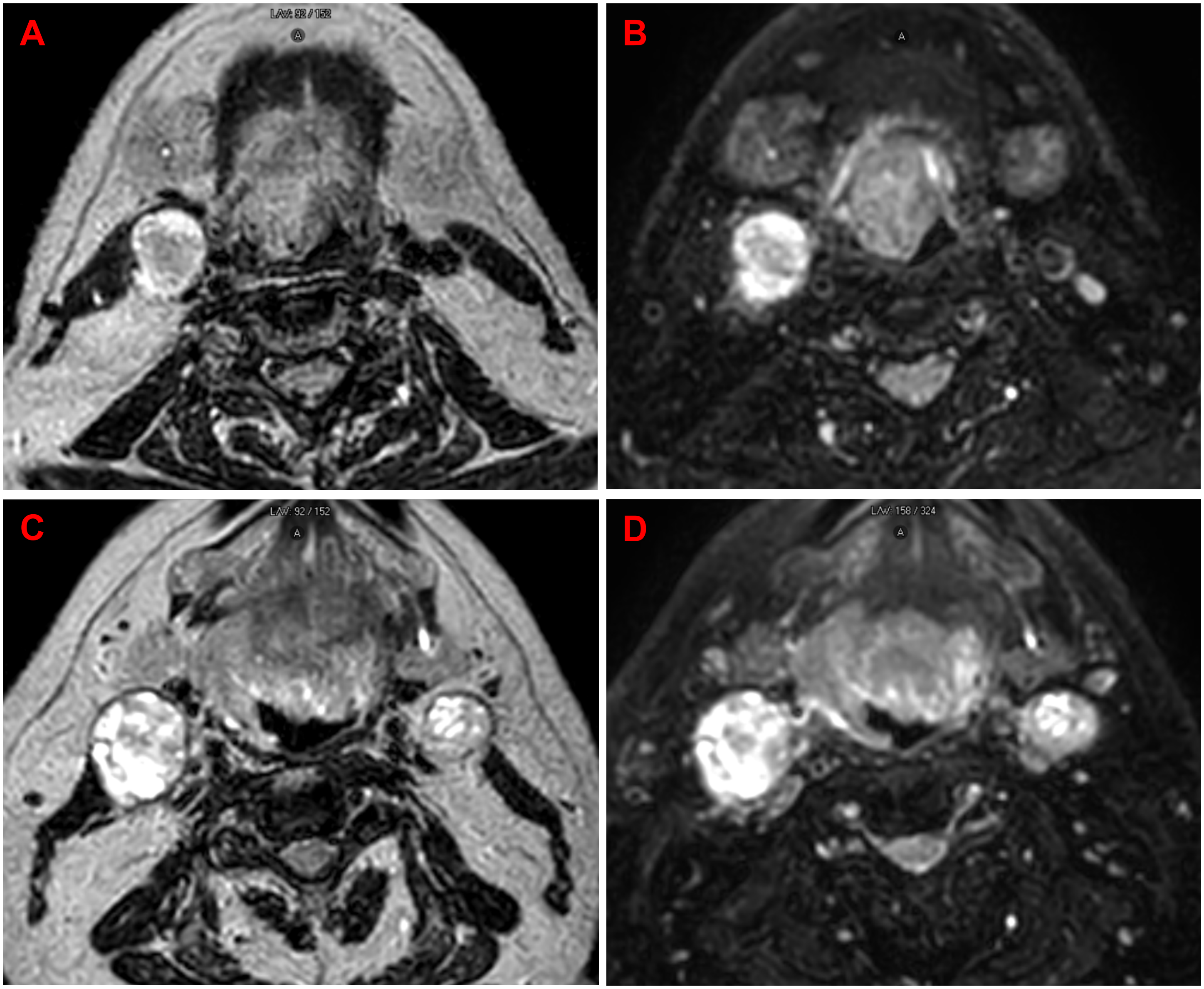


Figure S2: Representative cases of non-suppressed T2w (**A** and **C**) and SPAIR T2w (SPAIR 4) (**B** and **D**) in a HNC patient without visible segmentations. Refer to Figure 2 for segmentations annotating the primary GTV and metastatic lymph nodes.

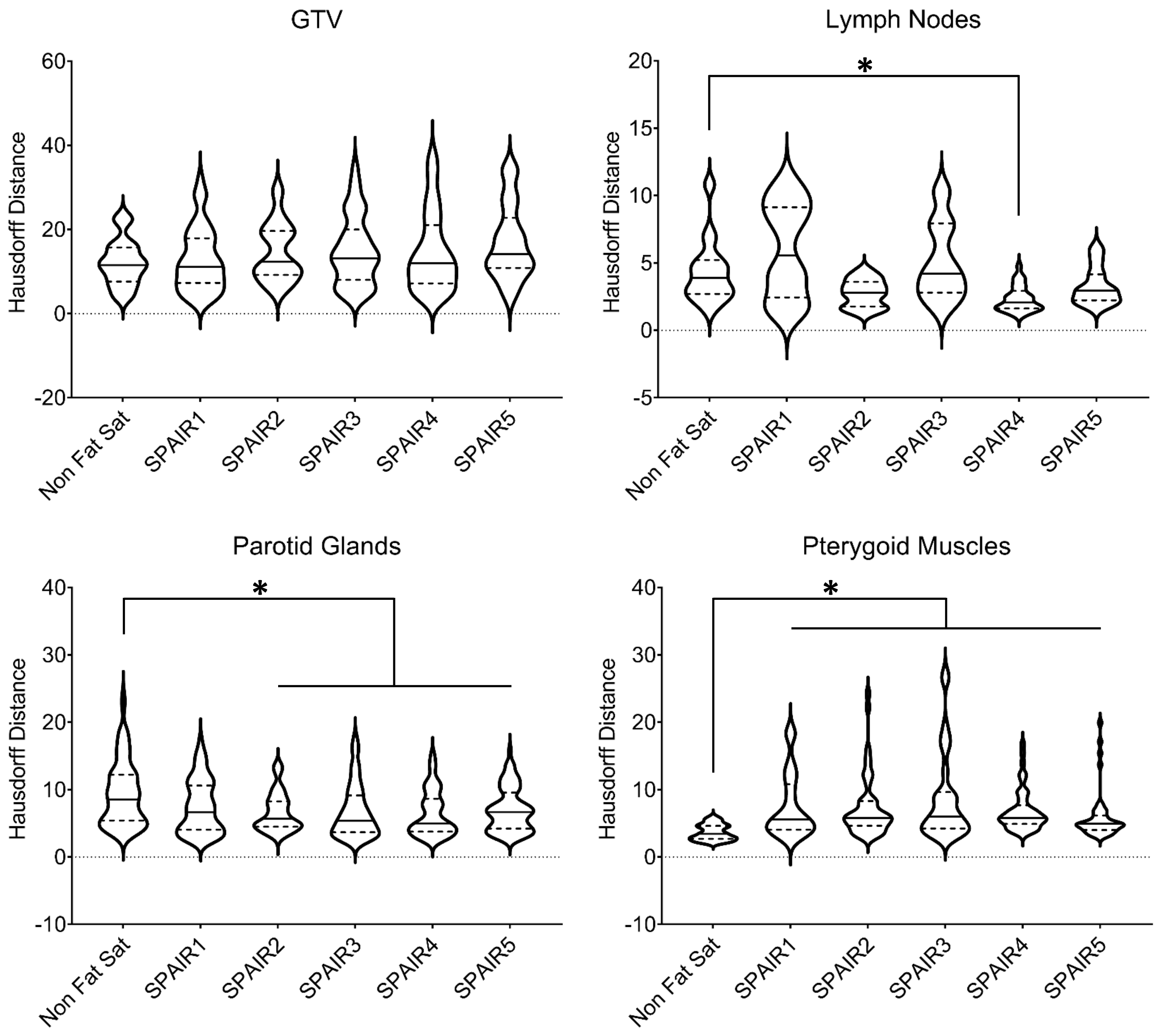


Figure S3: Hausdorff distance (HD) measurements of the GTV, lymph nodes, parotid glands, and pterygoid muscles in the non-suppressed and SPAIR sequences. Solid lines represent the median value of the distribution and dashed lines represent the limits of the interquartile range. * indicates significant differences (p<0.05). Although some of the violin plots extend below 0, all DSC values were positive.

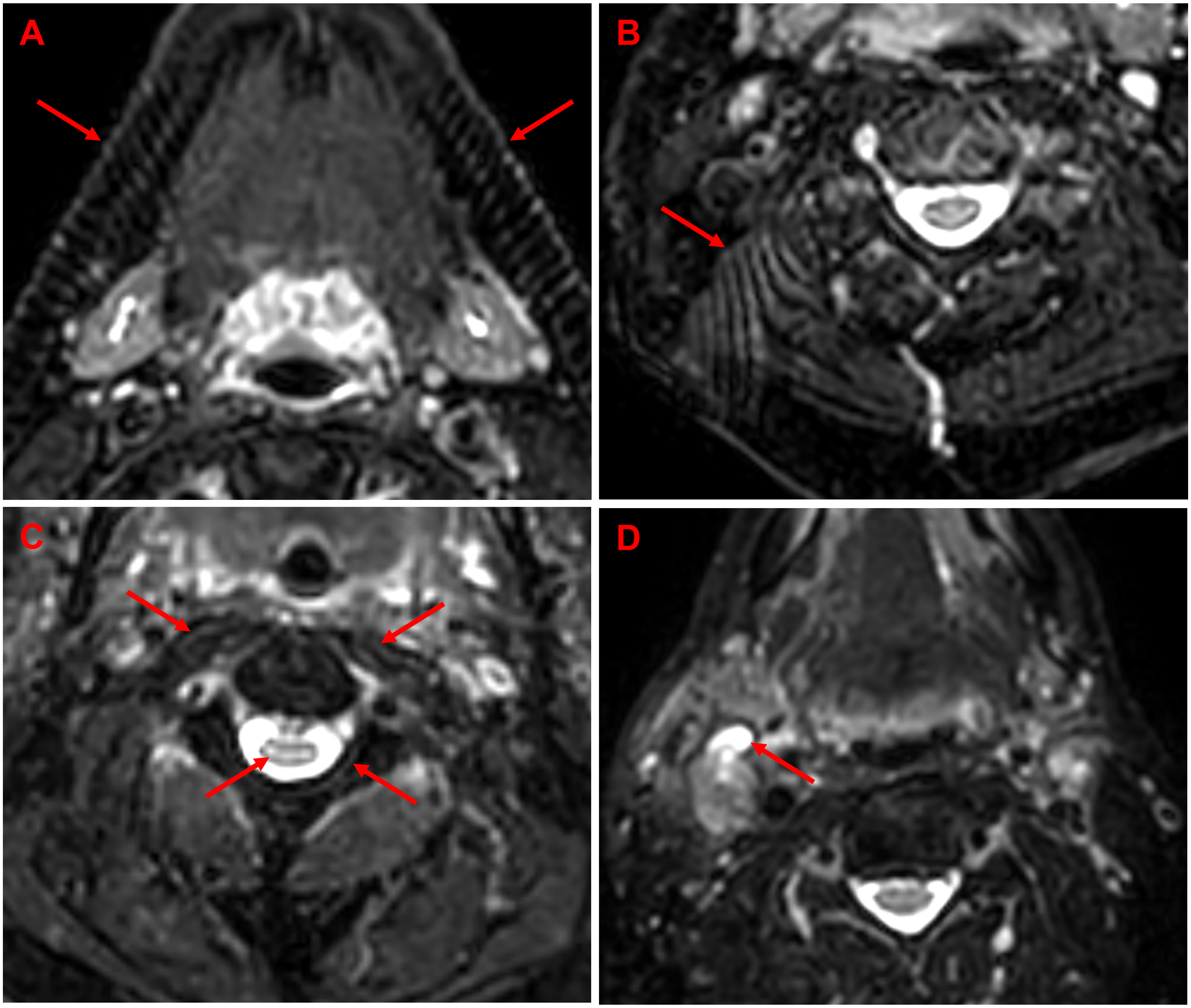


Figure S4: Representative examples of the artifacts that were observed in the SPAIR images. Herringbone (also called filtering) artifact is observed in the jaw in (**A**). Zebra artifact (also called 3D phase aliasing) is observed in the posterior area of the head (**B**). Gibb's ringing is observed around the spinal cord and other areas with abrupt intensity changes (**C**). A bright blood vessel is observed in (**D**), which is not observed on the contralateral side (this was the only patient in which this was observed).

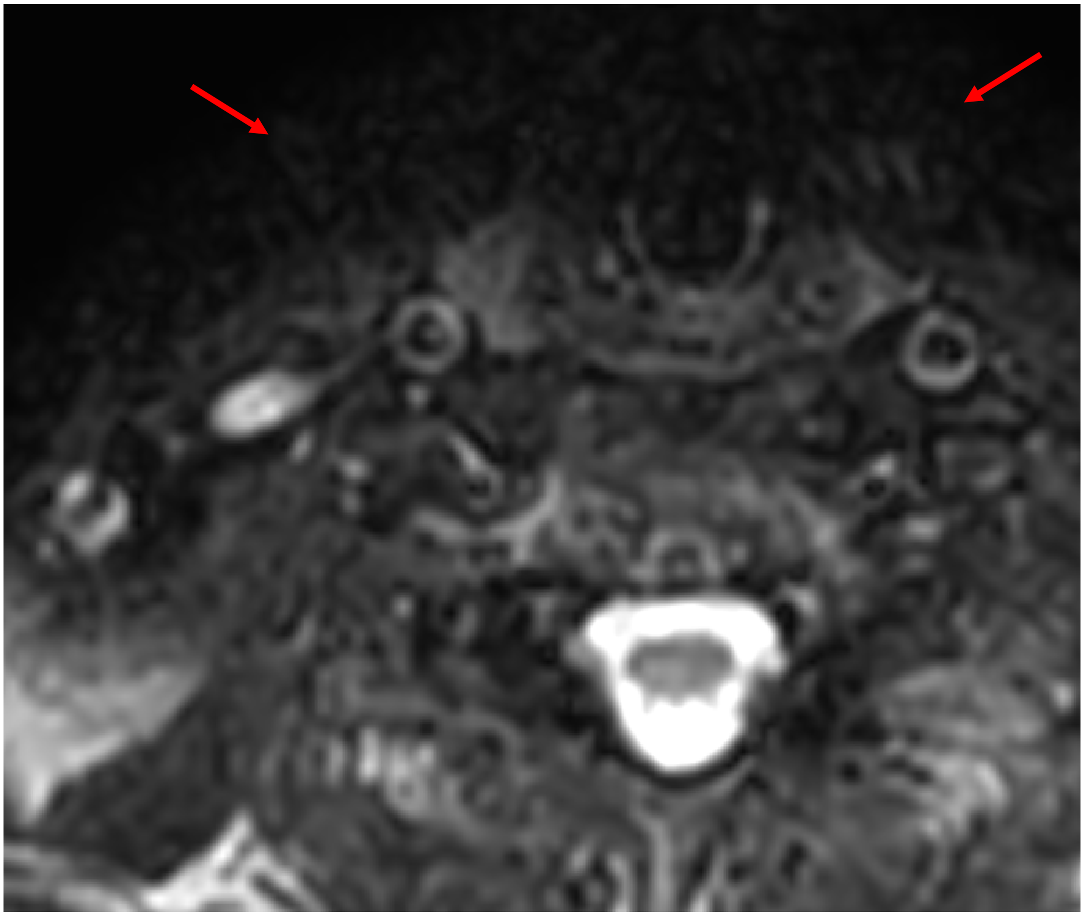

Figure S5: Example of burnout, which was observed in some capacity in each of the SPAIR sequences. This homogeneous hypointensity is a result of improper fat suppression in areas of B0 inhomogeneity. This is typically seen in the anterior and posterolateral areas of the image, which is far from isocenter and around tissue-air or tissue-bone interfaces.

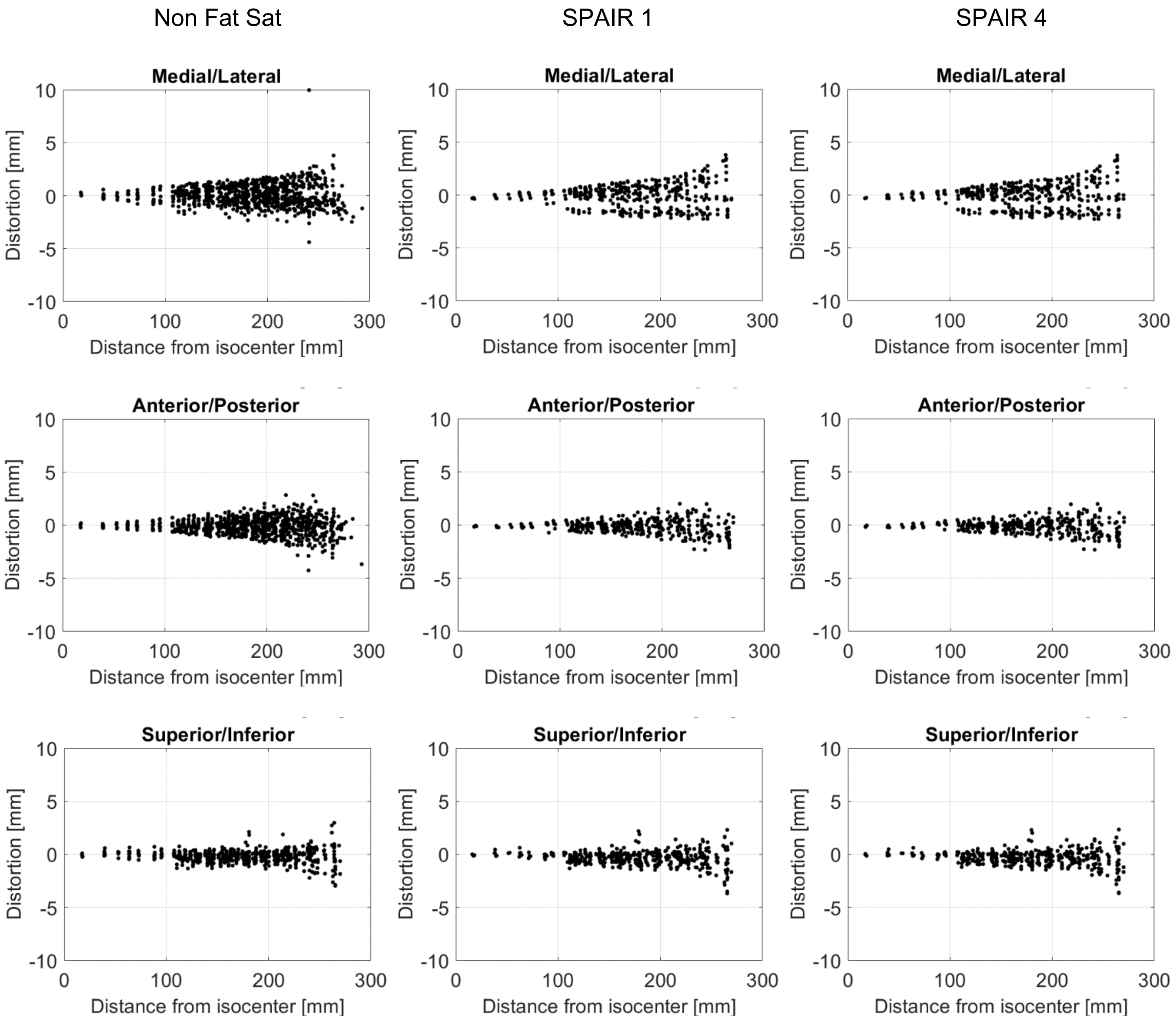


Figure S6: Geometric distortion distributions along the anatomic planes (rows) among the non-suppressed and SPAIR 1 and 4 sequences (columns). This data illustrates geometric distortion up to a diameter spherical volume (DSV) up to 500 mm, though the measurements in Table S6 are reflective of a DSV of 300 mm, since this is typically the maximum FOV used clinically. SPAIR 1 and 4 have very similar distributions, which is expected due to their very similar sequence parameters, but there are slight differences between the measurements, with a maximum discrepancy of 0.5 mm at one measurement location.
