## Appendix 1 for "Development and implementation of optimized endogenous contrast sequences for delineation in adaptive radiotherapy on a 1.5T MR-Linear-accelerator (MR-Linac): A prospective R-IDEAL Stage 0-2a quantitative/qualitative evaluation of *in vivo* site-specific quality-assurance using a 3D T2 fat-suppresse"

Appendix 1: Segmentor Analysis

### Instructions for Contouring MRI Cases

You will be assigned a Case Number, which you should use for each patient.

All cases are located in Raystation (Research).

At the bottom, you will see the individual MRI sequences which you can scroll through using the bar below. Double-click on the sequence that you want to work on. The first sequence is the non-fat-suppressed T2 image. The remaining images are the various iterations of fat-suppressed T2 images in random order. Double-clicking on one of the image viewers will maximize the window of the particular image plane. While you should use the Transverse plane to contour, you can refer to the other image planes to assist with visualization. When you are ready to contour, click the “New ROI geometry” icon on the header and select “Create new ROI”. Name the contour “XXX_YY” where XXX is the name of the organ and YY are your initials. “Organ” can remain as the “Type” and choose the color of your liking.

The organs that should be contoured are the:

- Tumor GTV
- Involved or suspicious lymph nodes
- L and R parotid glands
- L and R Pterygoid muscles

As we want to ensure that each observer’s contours are independent from each other, please do not look at other observer’s contours. If you cannot locate a particular organ, it would be best to ask someone not involved with the study (refer to initial email for those participating). Additionally, this study will assess the ability of each sequence to visualize these organs. Thus, please refrain from looking at the patient’s approved treatment plan. You may refer to any diagnostic exams and radiologist notes, as these are typically available to radiation oncologists during planning. Essentially, we would like to simulate the initial planning of a patient prescribed radiation. Time-permitting, please create each contour from scratch for each image, rather than propagating the contours between images of a patient.

Please see the next page for qualitative image assessment.

Qualitative Image Assessment

For each patient, rank each MR image according to preference for treatment planning (1 as lowest preference and 6 as highest preference). If you have the same preference for multiple images, give them the same rank. Provide the "Series description" which can be found by hovering your mouse over each image at the bottom of the screen.

| MRN | MR1 | MR2 | MR3 | MR4 | MR5 | MR6 |
|---|---|---|---|---|---|---|
| | Description:<br><br>Rank: | Description:<br><br>Rank: | Description:<br><br>Rank: | Description:<br><br>Rank: | Description:<br><br>Rank: | Description:<br><br>Rank: |
| | Description:<br><br>Rank: | Description:<br><br>Rank: | Description:<br><br>Rank: | Description:<br><br>Rank: | Description:<br><br>Rank: | Description:<br><br>Rank: |
| | Description:<br><br>Rank: | Description:<br><br>Rank: | Description:<br><br>Rank: | Description:<br><br>Rank: | Description:<br><br>Rank: | Description:<br><br>Rank: |
| | Description:<br><br>Rank: | Description:<br><br>Rank: | Description:<br><br>Rank: | Description:<br><br>Rank: | Description:<br><br>Rank: | Description:<br><br>Rank: |
| | Description:<br><br>Rank: | Description:<br><br>Rank: | Description:<br><br>Rank: | Description:<br><br>Rank: | Description:<br><br>Rank: | Description:<br><br>Rank: |

If you would like to elaborate on why an image was ranked high or low (such as presence of any artifacts or distortion), please list below. Provide the MRN and MR# with your finding.

Observer 1

1 = least preferred, 6 = most preferred

| MRN | MR1 | MR2 | MR3 | MR4 | MR5 | MR6 |
|---|---|---|---|---|---|---|
| Pat1 | Description: T2 3D tra<br><br>Rank: 5 | Description: SPAIR1<br><br>Rank: 6 | Description: SPAIR2<br><br>Rank: 3 | Description: SPAIR3<br><br>Rank:1 | Description: SPAIR4<br><br>Rank: 4 | Description: SPAIR5<br><br>Rank: 2 |
| Pat2 | Description: T2 3D tra<br><br>Rank: 6 | Description: SPAIR4<br><br>Rank: 2 | Description: SPAIR2<br><br>Rank3 | Description: SPAIR3<br><br>Rank:1 | Description: SPAIR5<br><br>Rank: 5 | Description: SPAIR1<br><br>Rank: 4 |
| Pat3 | Description: T2 3D tra<br><br>Rank: 2 | Description: SPAIR1<br><br>Rank: 4 | Description: SPAIR4<br><br>Rank: 5 | Description: SPAIR2<br><br>Rank: 3 | Description: SPAIR3<br><br>Rank: 6 | Description: SPAIR5<br><br>Rank: 1 |
| Pat4 | Description: T2 3D tra<br><br>Rank: 1 | Description: SPAIR1<br><br>Rank: 6 | Description: SPAIR2<br><br>Rank: 2 | Description: SPAIR4<br><br>Rank: 4 | Description: SPAIR5<br><br>Rank: 3 | Description: SPAIR3<br><br>Rank: 5 |
| Pat5 | Description: T2 3D tra<br><br>Rank: 1 | Description: SPAIR1<br><br>Rank: 4 | Description: SPAIR5<br><br>Rank: 6 | Description: SPAIR4<br><br>Rank: 2 | Description: SPAIR3<br><br>Rank: 5 | Description: SPAIR2<br><br>Rank:3 |

Other comments:

Pat3 SPAIR1: impossible to see left pterygoid. Had to guess

Pat3 SPAIR5: couldn’t see the GTV at all

Pat4 T2 3D tra: good for normals, harder for GTV

Pat1 SPAIR2: really hard to see everything. Very fuzzy. Lymph node was impossible to see

Pat1 SPAIR5: difficult to make out left parotid

Pat2 SPAIR4: impossible to see GTV

Pat2 SPAIR2: very difficult to see GTV

Pat2 SPAIR3: very fuzzy

Pat2 SPAIR1: really hard to make out parotids

Pat5 SPAIR1: GTV and parotids great, very hard to see pterygoids

Observer 2

1 = least preferred, 6 = most preferred

| MRN | MR1 | MR2 | MR3 | MR4 | MR5 | MR6 |
|---|---|---|---|---|---|---|
| Pat1 | Description:<br>T2 3D Tra<br><br>Rank: 5 | Description:<br>3D T2 SPAIR1<br><br>Rank: 6 | Description:<br>3D T2 SPAIR2<br><br>Rank: 5 | Description:<br>3D T2 SPAIR3<br><br>Rank: 1 | Description:<br>3D T2 SPAIR4<br><br>Rank: 5 | Description:<br>3D T2 SPAIR5<br><br>Rank: 5 |
| Pat2 | Description:<br>T2 3D Tra<br><br>Rank: 6 | Description:<br>3D T2 SPAIR4<br><br>Rank: 2 | Description:<br>3D T2 SPAIR2<br><br>Rank:3 | Description:<br>3D T2 SPAIR3<br><br>Rank: 5 | Description:<br>3D T2 SPAIR5<br><br>Rank:4 | Description:<br>3D T2 SPAIR1<br><br>Rank:5 |
| Pat3 | Description:<br>T2 3D Tra<br><br>Rank: 5 | Description:<br>3D T2 SPAIR1<br><br>Rank: 2 | Description:<br>3D T2 SPAIR4<br><br>Rank: 3 | Description:<br>3D T2 SPAIR2<br><br>Rank: 3 | Description:<br>3D T2 SPAIR3<br><br>Rank: 2 | Description:<br>3D T2 SPAIR5<br><br>Rank: 3 |
| Pat4 | Description:<br>T2 3D Tra<br><br>Rank: 6 | Description:<br>3D T2 SPAIR1<br><br>Rank: 4 | Description:<br>3D T2 SPAIR2<br><br>Rank: 3 | Description:<br>3D T2 SPAIR4<br><br>Rank: 4 | Description:<br>3D T2 SPAIR5<br><br>Rank: 5 | Description:<br>3D T2 SPAIR3<br><br>Rank: 4 |
| Pat5 | Description:<br>T2 3D Tra<br><br>Rank: 4 | Description:<br>3D T2 SPAIR1<br><br>Rank: 3 | Description:<br>3D T2 SPAIR5<br><br>Rank: 2 | Description:<br>3D T2 SPAIR4<br><br>Rank: 4 | Description:<br>3D T2 SPAIR3<br><br>Rank: 4 | Description:<br>3D T2 SPAIR2<br><br>Rank: 5 |

Observer 3

1 = least preferred, 6 = most preferred

| MRN | MR1 | MR2 | MR3 | MR4 | MR5 | MR6 |
|---|---|---|---|---|---|---|
| Pat1 | Description: GTV and LNs hard. Parotids very hard. Pterygoids good.<br><br>Rank: 1 | Description: GTV ok. LN hard. Parotids hard. Pterygoids hard.<br><br>Rank: 2 | Description: GTV ok. LN hard. Parotids ok. Pterygoids good.<br><br>Rank: 3 | Description: GTV ok. LN ok. Parotids good. Pterygoids ok.<br><br>Rank: 6 | Description: GTV ok. LN hard. Parotids good. Pterygoids ok.<br><br>Rank: 4 | Description: GTV ok. LN hard. Parotids good. Pterygoids good.<br><br>Rank: 5 |
| Pat2 | Description: GTV hard. LNs hard. Parotids hard. Pterygoids good.<br><br>Rank: 1 | Description: GTV and LNs good. Parotids and pterygoids good.<br><br>Rank: 6 | Description: GTV and LNs good. Parotids and pterygoids fair.<br><br>Rank: 4 | Description: GTV and LNs fair. Parotids and pterygoids good.<br><br>Rank: 5 | Description: GTV and LNs fair. Parotids and pterygoids good.<br><br>Rank: 3 | Description: GTV fair. LNs hard. Parotids good. Pterygoids fair.<br><br>Rank: 2 |
| Pat3 | Description: Pterygoids good. All other structures very hard.<br><br>Rank: 1 | Description: GTV and LN good. Pterygoids fair. Parotids hard.<br><br>Rank: 2 | Description: GTV and LN good. Pterygoids and parotids fair.<br><br>Rank: 5 | Description: GTV was fair. LN was great. Parotids and pterygoids were hard.<br><br>Rank: 3 | Description: GTV good. LN great. Parotids and pterygoids fair.<br><br>Rank: 6 | Description: GTV was fair. LN was good. Parotid and Pteryoids were fair.<br><br>Rank: 4 |
| Pat4 | Description: Really hard to see GTV. All other structures hard.<br><br>Rank: 1 | Description: GTV fair. Parotids and pterygoids hard.<br><br>Rank: 2 | Description: Fair GTV. Parotids hard. Pterygoids really hard.<br><br>Rank: 3 | Description: All structures are good.<br><br>Rank: 4 | Description: Best sequence for all structures.<br><br>Rank: 6 | Description: GTV is the best. Pterygoids and parotids are good.<br><br>Rank: 5 |
| Pat5 | Description: GTV and LN hard. Parotids and pterygoids good.<br><br>Rank: 1 | Description: GTV and LN good. Parotids and pterygoids hard.<br><br>Rank: 2 | Description: GTV and LN good. Parotids and pterygoids hard.<br><br>Rank: 4 | Description: GTV and LN good. Parotids and pterygoids the best.<br><br>Rank: 6 | Description: GTV and LN good. Parotids and pterygoids fair.<br><br>Rank: 3 | Description: GTV and LN good. Parotids great and pterygoids fair.<br><br>Rank: 5 |

Other comments:

Structures were hard to contour when the tissue around the structure had the same texture and contrast. Enhancement made the GTV and LNs easier. Pat1, Pat2: GTV and LNs were difficult in these cases. Pat3: artifact made the L pterygoid hard for this case.

Notes: (very) good = positive; (very) hard = negative; fair/okay = neutral

Observer 4

1 = least preferred, 6 = most preferred

| MRN | MR1 | MR2 | MR3 | MR4 | MR5 | MR6 |
|---|---|---|---|---|---|---|
| Pat1 | Description: T2 3D Tra<br><br>Can't see parotid<br>Easier to see LN remnant<br>Had to imagine parts of GTV<br><br>Rank: 4 | Description: SPAIR1<br><br>Can't see LN remnant<br>Easiest to see GTV<br><br>Rank: 6 | Description: SPAIR2<br><br>Can't see LN remnant, had to hypothetically create<br>Second easiest to see GTV; easiest for parotid<br><br>Rank: 5 | Description: SPAIR3<br><br>Can't see LN remnant, had to hypothetically create<br><br>Rank: 3 | Description: SPAIR4<br><br>Can't see LN remnant, had to hypothetically create<br><br>Rank: 2 | Description: SPAIR5<br><br>Can't see LN remnant, had to hypothetically create<br>Had to imagine parts of GTV<br><br>Rank: 1 |
| Pat2 | Description: T2 3D Tra<br><br>Had to guess parotid gland; had to imagine GTV<br><br>Rank: 1 | Description: SPAIR4<br><br>Fourth easiest to see GTV<br><br>Rank: 3 | Description: SPAIR2<br><br>Easiest to see parotid gland<br>Second easiest to see GTV<br><br>Rank: 5 | Description: SPAIR3<br><br>Third easiest to see GTV<br><br>Rank: 4 | Description: SPAIR5<br><br>Difficult to see GTV<br><br>Rank: 2 | Description: SPAIR1<br><br>Easiest to see GTV<br><br>Rank: 6 |
| Pat3 | Description: T2 3D Tra<br><br>Impossible to see parotids and lymph nodes; had to guess to see them<br>hard to see GTV<br><br>Rank: 1 | Description: SPAIR1<br><br>Second easiest to see parotids<br>Easiest to see GTV<br><br>Rank: 6 | Description: SPAIR4<br><br>Easiest to see parotid gland<br>Second easiest to see GTV<br><br>Rank: 5 | Description: SPAIR2<br><br>Moderately easy to see GTV, but not as easily as base scan or 20201104<br><br>Rank: 4 | Description: SPAIR3<br><br>Reasonably good series<br><br>Rank: 3 | Description: SPAIR5<br><br>most challenging to see GTV<br><br>Rank: 2 |
| Pat4 | Description: T2 3D Tra<br><br>Very hard to see parotid glands<br><br>Rank: 1 | Description: SPAIR1<br><br>Challenging to see pterygoids fully; easiest to see parotids<br>Third easiest to see GTV<br><br>Rank: 4 | Description: SPAIR2<br><br>Harder to see parotids than others<br>Easiest to see GTV<br><br>Rank: 6 | Description: SPAIR4<br><br>Very blurry<br>Third easiest to see GTV<br><br>Rank: 3 | Description: SPAIR5<br><br>Easiest to see parotid gland<br>Second easiest to see GTV<br><br>Rank: 5 | Description: SPAIR3<br><br>Challenging to see parotids<br><br>Rank: 2 |
| Pat5 | Description: T2 3D Tra<br><br>Impossible to see parotids; had to guess to see them<br>Impossible to see LN<br><br>Rank: 1 | Description: SPAIR1<br><br>Easiest to see GTV<br><br>Rank: 6 | Description: SPAIR5<br><br>So blurry<br>Reasonably easy to see GTV<br><br>Rank: 2 | Description: SPAIR4<br><br>Very blurry<br>Third easiest to see GTV<br><br>Rank: 3 | Description: SPAIR3<br><br>Reasonably good series<br><br>Rank: 4 | Description: SPAIR2<br><br>Second easiest to see GTV<br><br>Rank: 5 |

Notes: 5$^{th}$ and 6$^{th}$ easiest = negative; 1$^{st}$ and 2$^{nd}$ easiest = positive; 3$^{rd}$ and 4$^{th}$ easiest = neutral

Observer 5

1 = least preferred, 6 = most preferred

| MRN | MR1 | MR2 | MR3 | MR4 | MR5 | MR6 |
|---|---|---|---|---|---|---|
| Pat 1 | Description: T2 3D Tra<br><br>Rank: 1 | Description: 3D T2 SPAIR1<br><br>Rank: 6 | Description: 3D T2 SPAIR2<br><br>Rank: 4 | Description: 3D T2 SPAIR3<br><br>Rank: 5 | Description: 3D T2 SPAIR4<br><br>Rank: 3 | Description: 3D T2 SPAIR5<br><br>Rank: 2 |
| Pat2 | Description: T2 3D Tra<br><br>Rank: 1 | Description: 3D T2 SPAIR4<br><br>Rank: 6 | Description: 3D T2 SPAIR2<br><br>Rank: 5 | Description: 3D T2 SPAIR3<br><br>Rank: 4 | Description: 3D T2 SPAIR5<br><br>Rank: 3 | Description: 3D T2 SPAIR1<br><br>Rank: 2 |
| Pat3 | Description: T2 3D Tra<br><br>Rank: 1 | Description: 3D T2 SPAIR1<br><br>Rank: 6 | Description: 3D T2 SPAIR4<br><br>Rank: 4 | Description: 3D T2 SPAIR2<br><br>Rank: 3 | Description: 3D T2 SPAIR3<br><br>Rank: 5 | Description: 3D T2 SPAIR5<br><br>Rank: 2 |
| Pat4 | Description: T2 3D Tra<br><br>Rank: 1 | Description: 3D T2 SPAIR1<br><br>Rank: 2 | Description: 3D T2 SPAIR2<br><br>Rank: 4 | Description: 3D T2 SPAIR4<br><br>Rank: 3 | Description: 3D T2 SPAIR5<br><br>Rank: 5 | Description: 3D T2 SPAIR3<br><br>Rank: 6 |
| Pat5 | Description: T23D Tra<br><br>Rank: 1 | Description: 3D T2 SPAIR1<br><br>Rank: 6 | Description: 3D T2 SPAIR5<br><br>Rank: 5 | Description: 3D T2 SPAIR4<br><br>Rank: 4 | Description: 3D T2 SPAIR3<br><br>Rank: 3 | Description: 3D T2 SPAIR2<br><br>Rank: 2 |

Other comments:

For all T2 3D Tra: the muscles were well visualized but all other structures were significantly more difficult to evaluate.
