## Appendix 2 for "Development and implementation of optimized endogenous contrast sequences for delineation in adaptive radiotherapy on a 1.5T MR-Linear-accelerator (MR-Linac): A prospective R-IDEAL Stage 0-2a quantitative/qualitative evaluation of *in vivo* site-specific quality-assurance using a 3D T2 fat-suppresse"

### Appendix 2: MR Physicist Analysis

#### SPAIR Image Quality Assessment

There are 5 SPAIR T2 images and 1 non-fat-suppressed T2 image for each patient. While looking through each image, please fill out the chart on the following pages with information about the image quality and presence of artifacts. Regarding the chart:

- List the series name/description (T2, SPAIR1, SPAIR2, SPAIR3, SPAIR4, SPAIR5)
- List the level of fat suppression (low, moderate, high)
- List any artifacts and provide the severity of the artifact (low, moderate, high)
- Provide any additional notes you may have about the image quality (i.e. clarity, qualitative contrast, etc.)
- For the SPAIR images (excluding the non-fat-suppressed T2), provide the relative rank of the overall image quality considering all artifacts and level of fat suppression
    - 1 for worst image quality, 5 for the best image quality

MR Physicist 1

| MRN: Pat1 | | | |
|---|---|---|---|
| Series Name: | Image Quality Assessment | Other Notes | Rank |
| T2 Tra (MRTC 6 min) | Fat Suppression: N/A<br><br>Artifacts (list and provide severity)<br>Minor inhomogeneity artifacts (hyperintensity/dropout) near dental metal and IV port on chest | Without fat sat, more difficult to determine gland boundaries & harder to distinguish gland from muscle based on signal intensity | |
| 3D T2 SPAIR5 | Fat Suppression: Modest, anterior burnout on lowest 70 slices of 250<br><br>Artifacts (list and provide severity); herringbone artifact and some Gibbs ringing near muscle and gland structures, 2 severity | | 2 |
| 3D T2 SPAIR1 | Fat Suppression: Good posteriorly from roof of nasal cavity down to C4, good anteriorly from top slice to C5, then some burnout of anterior skin until C7, then no fat sat<br><br>Artifacts (list and provide severity) burnout (water saturation) anterior to mouth due to susceptibility artifact from positioning device | Easy to distinguish glands & other structures. Skin contour preserved for most of the image stack.<br>Images appear a little smooth | 3 |
| 3D T2 SPAIR2 | Fat Suppression:<br>Good overall, mild burnout in anterior only for lowest 35 slices, signal loss near positioning device<br><br>Artifacts (list and provide severity):<br>Herringbone artifact (fine lines) on both sides of jaw, severity 3, would interfere with auto segmentation. Ringing (Gibbs artifact) seen near muscle/fat interfaces, lowers apparent resolution, severity 2 | Gland conspicuity is good, but ringing and blurriness make this more difficult to enjoy. | 2 |
| 3D T2 SPAIR3 | Fat Suppression: Good overall, mild burnout in anterior only for lowest 35 slices, mild signal loss near positioning device<br><br>Artifacts (list and provide severity): No obvious ringing or Gibbs artifact, | Good differentiation between gland, muscle and fat<br>Image sharpness looks good | 5 |

| | | | |
|---|---|---|---|
| 3D T2 SPAIR4 | Fat Suppression: Very good. mild burnout in anterior only for lowest 35 slices, mild signal loss near positioning device<br><br>Artifacts (list and provide severity): No obvious ones except signal loss | Good differentiation between gland, muscle and fat<br>Image sharpness looks good, maybe not quite as good as SPAIR3 | 4 |

| MRN: Pat2 | | | |
|---|---|---|---|
| Series Name: | Image Quality Assessment | Other Notes | Rank |
| T2 Tra | Fat Suppression: N/A<br><br>Artifacts (list and provide severity): signal dropouts near immobilization device | SNR high, conspicuity of glands and tumor lower<br>Apparent metal artifact or inhomogeneity near right shoulder causing artifacts in all scans | |
| 3D T2 SPAIR4 | Fat Suppression: Inhomogeneous, burnout posteriorly from slices 80-130, no fat sat posteriorly below slice 80 or above slice 150<br><br>Artifacts (list and provide severity): Bright vessels and some blurriness of glands/tumor, severity 3 (moderate). | Glands visible but bright vascular structures nearby may limit segmentation | 5 |
| 3D T2 SPAIR2 | Fat Suppression: Signal burnout anterior up to slice 80 and posterior slices 100-150<br><br>Artifacts (list and provide severity): flame artifact (zebra pattern) slices 120-140 posterior right side, severity 2 because not near tumor but would be higher if covering anatomy of interest; herringbone artifact near jaw, severity low (1) | Some blurriness in tumor & gland depiction | 2 |
| 3D T2 SPAIR3 | Fat Suppression: Aggressive<br>Anterior burnout to slice 80, lateral posterior burnout (loss of skin contour) slices 110-145<br><br>Artifacts (list and provide severity): | Gland and tumor conspicuity are good | 3 |
| 3D T2 SPAIR5 | Fat Suppression:<br>Anterior burnout to slice 100, posterolateral burnout slices 120-150<br><br>Artifacts (list and provide severity): herringbone on both sides of jaw, severity 2, medium, some Gibbs ringing near structures. | Some blurriness in gland depiction | 1 |

| | | | |
|---|---|---|---|
| 3D T2 SPAIR1 | Fat Suppression:<br>Anterior burnout to slice 90 including trachea from slices 50-80<br>Posterolateral burnout slices 110-145<br><br>Artifacts (list and provide severity): | Bright vascular structures obscure some boundaries of glands & tumor | 3 |

| MRN: Pat3 | | | |
|---|---|---|---|
| Series Name: | Image Quality Assessment | Other Notes | Rank |
| 3D T2 | Fat Suppression: N/A<br><br>Artifacts (list and provide severity): significant signal dropout due to dental metal | Tumor conspicuity is low | |
| 3D T2 SPAIR1 | Fat Suppression:<br>Anterior burnout from slice 30-80<br><br>Artifacts (list and provide severity): loss of fat sat near dental metal | Gland conspicuity is still good even where fat sat is failing due to metal | 4 |
| 3D T2 SPAIR4 | Fat Suppression: Very inhomogeneous. Burnout anteriorly from slice 30-100, also signal dropoff at lateral shoulders at bottom of imaging stack<br><br>Artifacts (list and provide severity): loss of fat sat and signal near dental metal, obscuring anatomy, high severity (3? 5?) | Gland boundary almost obscured by fat sat artifacts | 2 |
| 3D T2 SPAIR2 | Fat Suppression:<br>Anterior burnout from slice 20-90<br><br>Artifacts (list and provide severity):<br>flame artifact (zebra pattern) slices 110-140 posterior right side, severity 2 because not near tumor but would be higher if covering anatomy of interest; loss of fat sat and signal near dental metal, obscuring anatomy, high severity (3? 5?) | Gland boundary almost obscured by fat sat artifacts | 1 |
| 3D T2 SPAIR3 | Fat Suppression:<br>Anterior burnout from slice 30-80<br><br>Artifacts (list and provide severity): loss of fat sat and signal near dental metal, obscuring anatomy, high severity (3? 5?) | Gland boundary almost obscured by fat sat artifacts | 5 |

| | | | |
|---|---|---|---|
| 3D T2 SPAIR5 | Fat Suppression: Inhomogenous; Anterior burnout slices 60-<br>100<br><br>Artifacts (list and provide severity): Respiratory artifact due to lower placement of imaging stack obscures chest wall from slices 1-60 ; herringbone artifact on anterior cheeks slices 120-180 low severity. | Gland boundary obscured by fat sat failure | 2 |

| MRN: Pat4 | | | |
|---|---|---|---|
| Series Name: | Image Quality Assessment | Other Notes | Rank |
| T2 3D Tra | Fat Suppression:N/A<br><br>Artifacts (list and provide severity) | Gland and tumor conspicuity are low | |
| 3D T2 SPAIR1 | Fat Suppression:<br>Burnout posteriorly slices 180-130 then losing fat sat posteriorly below slice 130<br><br>Artifacts (list and provide severity): | Good parotid gland conspicuity, lower contrast for some other structures near muscle. (Image looks a little flat.) | 4 |
| 3D T2 SPAIR2 | Fat Suppression:<br>Posterior burnout from slice 130-190, no posterior fat sat below slice 130<br><br>Artifacts (list and provide severity): flame artifact posteriorly slices 147-114 severity 2.; herringbone/ringing artifact slices 180-130 obscuring gland interfaces, severity high (3) | Gland edges would be disrupted by artifact | 1 |
| 3D T2 SPAIR4 | Fat Suppression:<br>Anterior burnout slice 5-75, posterior burnout slices 84-120<br><br>Artifacts (list and provide severity) | Gland conspicuity good, no apparent artifacts; even a nice view of the optic nerves with fat sat near slice 200 | 5 |
| 3D T2 SPAIR5 | Fat Suppression:<br>Posterior burnout slice 150-205<br>Anterior burnout or indistinct slices 85-145<br><br>Artifacts (list and provide severity): herringbone artifacts slices 150-180 severity 1 but adjacent to glands | Good aortic arch depiction (irrelevant but nice) | 2 |

| | | | |
|---|---|---|---|
| 3D T2 SPAIR3 | Fat Suppression:<br>Anterior burnout slices 60-130<br>Posterior burnout from 200 down to 135 where fat sat fails<br><br>Artifacts (list and provide severity) | Mostly dark vessels help visualization | 3 |

| MRN: Pat5 | | | |
|---|---|---|---|
| Series Name: | Image Quality Assessment | Other Notes | Rank |
| T2 3D Tra | Fat Suppression: N/A<br><br>Artifacts (list and provide severity) | Minor artifact from dental metal | |
| 3D T2 SPAIR1 | Fat Suppression:<br>Anterior burnout slices 20-75<br>Posterior burnout from slices 100-140<br><br>Artifacts (list and provide severity) | A little blurry | 2 |
| 3D SPAIR5 | Fat Suppression:<br>Anterior burnout slice 60-95, not as severe as usual<br><br>Artifacts (list and provide severity): herringbone on cheeks and edges of glands severity 2. | Mild breakdown of fat sat near dental metal, not much anatomy obscured | 2 |
| 3D T2 SPAIR4 | Fat Suppression:<br>Anterior burnout slices 5-65<br>; posterior burnout slices 85-120<br><br>Artifacts (list and provide severity) | Good gland & tumor conspicuity | 5 |
| 3D T2 SPAIR3 | Fat Suppression:<br>Anterior burnout or missing contour from slice 1-55; Posterior burnout from slices 55-95<br><br>Artifacts (list and provide severity): low SNR posterior neck region | | 3 |

| | | | |
|---|---|---|---|
| 3D T2 SPAIR2 | Fat Suppresion:<br>Posterior burnout slices 55-100; anterior burnout slices 5-50 (bad burnout on slices 10-20)<br><br>Artifacts (list and provide severity):<br>Flame artifact posterior slices 65-85 | CNR very good (high conspicuity) | 3 |

MR Physicist 2

SPAIR Image Quality Assessment

| MRN: Pat1 | | | |
|---|---|---|---|
| Series Name: | Image Quality Assessment | Other Notes | Rank |
| SPAIR1 | Fat Suppression: moderate<br><br>Artifacts (list and provide severity): low | All comments for HN area only, not off isocenter. | 4 |
| SPAIR4 | Fat Suppression: high<br><br>Artifacts (list and provide severity): low | Higher T2 contrast | 5 |
| SPAIR2 | Fat Suppression: moderate<br><br>Artifacts (list and provide severity): filtering - moderate | *filtering – edge effects likely cause by echo sampling scheme. | 2 |
| SPAIR3 | Fat Suppression: moderate<br><br>Artifacts (list and provide severity): low | | 3 |

| | | | |
|---|---|---|---|
| SPAIR5 | Fat Suppression: moderate<br><br>Artifacts (list and provide severity):<br>filtering – moderate<br>partial volume – moderate<br>Due to noticeably thicker slices | | 1 |

| MRN: Pat2 | | | |
|---|---|---|---|
| Series Name: | Image Quality Assessment | Other Notes | Rank |
| SPAIR1 | Fat Suppression: moderate<br><br>Artifacts (list and provide severity): low | All comments for HN area only, not off isocenter. | 4 |
| SPAIR4 | Fat Suppression: high<br><br>Artifacts (list and provide severity): low | Higher T2 contrast | 5 |
| SPAIR2 | Fat Suppression: moderate<br><br>Artifacts (list and provide severity):<br>filtering – moderate | *filtering – edge effects likely cause by echo sampling scheme. | 2 |
| SPAIR3 | Fat Suppression: moderate<br><br>Artifacts (list and provide severity): low | | 3 |
| SPAIR5 | Fat Suppression: moderate<br><br>Artifacts (list and provide severity):<br>filtering – moderate<br>partial volume - moderate | | 1 |

| MRN: Pat3 | | | |
|---|---|---|---|
| Series Name: | Image Quality Assessment | Other Notes | Rank |
| SPAIR1 | Fat Suppression: moderate<br><br>Artifacts (list and provide severity):<br>Metal - moderate | All comments for HN area only, not off isocenter.<br><br>Failure around metal similar for all images | 4 |
| SPAIR4 | Fat Suppression: high<br><br>Artifacts (list and provide severity):<br>Metal - moderate | Higher T2 contrast | 5 |
| SPAIR2 | Fat Suppression: moderate<br><br>Artifacts (list and provide severity):<br>Metal - moderate<br>filtering – moderate<br>aliased phase - high | *filtering – edge effects likely cause by echo sampling scheme. | 1 |
| SPAIR3 | Fat Suppression: moderate<br><br>Artifacts (list and provide severity):<br>Metal - moderate | | 3 |
| SPAIR5 | Fat Suppression: moderate<br><br>Artifacts (list and provide severity):<br>Metal - moderate<br>filtering – moderate<br>partial volume - moderate | | 2 |

| MRN: Pat4 | | | |
|---|---|---|---|
| Series Name: | Image Quality Assessment | Other Notes | Rank |
| SPAIR1 | Fat Suppression: moderate<br><br>Artifacts (list and provide severity): low | All comments for HN area only, not off isocenter. | 4 |
| SPAIR4 | Fat Suppression: high<br><br>Artifacts (list and provide severity): low | Higher T2 contrast | 5 |
| SPAIR2 | Fat Suppression: moderate<br><br>Artifacts (list and provide severity):<br>filtering – moderate | *filtering – edge effects likely cause by echo sampling scheme. | 2 |
| SPAIR3 | Fat Suppression: moderate<br><br>Artifacts (list and provide severity): low | | 3 |
| SPAIR5 | Fat Suppression: moderate<br><br>Artifacts (list and provide severity):<br>filtering – moderate<br>partial volume - moderate | | 1 |

| MRN: Pat5 | | | |
|---|---|---|---|
| Series Name: | Image Quality Assessment | Other Notes | Rank |
| SPAIR1 | Fat Suppression: low<br><br>Artifacts (list and provide severity): low | All comments for HN area only, not off isocenter. | 3 |
| SPAIR4 | Fat Suppression: high<br><br>Artifacts (list and provide severity): low | Higher T2 contrast | 5 |
| SPAIR2 | Fat Suppression: moderate<br><br>Artifacts (list and provide severity):<br>filtering – moderate<br>aliased phase - moderate | *filtering – edge effects likely cause by echo sampling scheme. | 2 |
| SPAIR3 | Fat Suppression: moderate<br><br>Artifacts (list and provide severity): low | | 4 |
| SPAIR5 | Fat Suppression: moderate<br><br>Artifacts (list and provide severity):<br>filtering – moderate<br>partial volume - moderate | | 1 |
